# Thrifty energy phenotype predicts weight regain - results of a randomized controlled trial

**DOI:** 10.1101/2021.03.25.21254300

**Authors:** Leonard Spranger, Josephine Bredow, Ulrike Zeitz, Ulrike Grittner, Michael Boschmann, Sophia Dickmann, Nicole Stobäus, Reiner Jumpertz-von Schwartzenberg, Joachim Spranger, Knut Mai

**Affiliations:** Department of Endocrinology and Metabolism, Charité - Universitätsmedizin Berlin, corporate member of Freie Universität Berlin, Humboldt-Universität zu Berlin, and Berlin Insitute of Health, 10117 Berlin, Germany; Charité Center for Cardiovascular Research, Charité – Universitätsmedizin Berlin, corporate member of Freie Universität Berlin, Humboldt-Universität zu Berlin, and Berlin Institute of Health, 10117 Berlin, Germany; DZHK (German Centre for Cardiovascular Research), partner site Berlin; Institute of Biometry and Clinical Epidemiology, Charité - Universitätsmedizin Berlin, corporate member of Freie Universität Berlin, Humboldt-Universität zu Berlin, and Berlin Institute of Health, Berlin, Germany; Berlin Institute of Health (BIH), Anna-Louisa-Karsch-Str. 2, 10178 Berlin, Germany; Experimental and Clinical Research Center (ECRC) - Charité - Universitätsmedizin Berlin and Max Delbrueck Center for Molecular Medicine, Berlin, Germany; Clinical Research Unit, Charité – Universitätsmedizin Berlin, corporate member of Freie Universität Berlin, Humboldt-Universität zu Berlin, and Berlin Institute of Health, 10117 Berlin, Germany

**Keywords:** lean body mass, fat mass, insulin sensitivity, obesity, weight loss, energy restriction, energy metabolism

## Abstract

**Background & Aims:** Weight loss is associated with an improvement of insulin sensitivity. Both, a negative energy balance and changes of body composition are integrative components of weight loss interventions. However, the individual impact of these two components on insulin sensitivity and energy metabolism is unclear.

**Methods:** We performed a randomized controlled trial including 80 overweight or obese post-menopausal women. Participants randomly assigned to the intervention group underwent an 800 kcal/d liquid diet for 2 months followed by four weeks in which the formula diet was substituted by a calorie reduced healthy diet to facilitate further weight loss. This weight loss phase was followed by a 4-week weight maintenance phase, where weight stability was achieved by individualized daily caloric intake without negative energy balance. Volunteers of the control group were instructed to keep their weight stable during the entire period of 4 months. Metabolic phenotyping was performed in both groups at baseline (M0), after weight loss (M3) and after the maintenance period (M4). Additional phenotyping was performed during follow-up at 12 (M12) and 24 months (M24). Primary outcomes were changes of lean body mass (LBM) and changes of insulin sensitivity (ISI_Clamp_) between baseline and M3 and M4. Estimates of energy metabolism were secondary endpoints.

**Results:** No significant changes of body weight or LBM were found in the control group between any time points. A significant reduction of body weight, fat mass (FM) and LBM was found in the intervention group between M0 and M3, while no further change was seen between M3 and M4. Only subjects of the intervention group were characterized by an improvement of the second primary outcome ISI_Clamp_ at M3, which was preserved until M4. Notably, a lower resting energy expenditure per LBM (REE_LBM_) at M3 as well as the individual difference of REE_LBM_ between M3 and M4 significantly predicted a stronger regain of fat mass during follow-up.

**Conclusions:** In summary, our data demonstrate that modulation of LBM and insulin sensitivity during weight loss is predominantly driven by changes in body weight and body composition, rather than an individual effect of negative energy balance. However, the variance in energy expenditure during negative and steady energy balance indicates a thrifty phenotype, which is highly susceptible to future regain of fat mass.

## 1. Introduction

Obesity is a risk factor of numerous metabolic disorders. Correspondingly, weight loss is associated with an improvement of lipid and glucose metabolism as well as whole body and tissue specific insulin resistance. Changes of hormones relevant in energy homeostasis have been suggested to reflect a counter-regulatory neuro-endocrine response promoting subsequent weight regain^1-4^. Notably, short-term hypocaloric intervention can also partially reverse obesity related metabolic dysbalance^5-7^. Both, a negative energy balance and changes of body composition are integrative components of weight loss interventions and the specific individual impact of negative energy balance versus the effect of a changed body composition on changes of insulin sensitivity and energy metabolism were not yet differentiated. In fact, these effects could be synergistic, antagonistic or independent. A better understanding of this processes might support a prediction of the frequently observed weight regain following successful body weight reduction^8,9^ as weight loss induced hormonal changes have been suggested to be involved in this phenomenon. Accordingly, we recently demonstrated the impact of weight loss-induced changes of adrenergic activity and adipose tissue metabolism on body weight maintenance^2,10^. Although numerous other mechanisms have been suspected to be involved in body weight regain, reported data are frequently inconsistent. Exemplarily, divergent findings have been reported indicating positive, negative or no associations between leptin or ghrelin and weight regain^11-15^.

Different states of weight loss and energy deficiency may cause such conflicting data. Therefore, a detailed analyses and separation of negative energy balance and improved body weight and/or body composition is desirable. The identification of specific effects of energy balance and changed body composition/body weight on long-term regulation of obesity may improve our understanding of body weight regulation. As no or only temporary delay of subsequent weight regain was achieved by a prolonged weight maintenance intervention^9,10^, this knowledge should be helpful to improve current strategies focusing on sustained long-term weight reduction. Such strategies are highly desirable considering that a reduction of cardiovascular events could be achieved by sustained weight loss as demonstrated in patients successfully reducing body weight in the LookAHEAD trial^16^. An integral component of successful weight loss is the improvement of whole body and tissue specific insulin sensitivity. However, whether changes in body composition alone or only in combination with a negative energy balance determine the improvements in insulin sensitivity during weight loss remains unknown.

Therefore, we designed a randomized controlled weight loss intervention trial integrating a weight maintenance phase after weight loss to dissociate negative energy balance from changes in body composition. Our data indicate that improved insulin sensitivity and changes in LBM after weight loss are mainly driven by changes in body composition independent of a negative energy balance. Secondary analyses revealed that the individual response of energy expenditure to the negative energy balance during weight loss predicts future fat mass regain.

## 2. Materials and Methods

### Participants

Between March 2012 and July 2015 a total of 80 overweight or obese post-menopausal women were included in this study. The last volunteers finished the study in September 2017, when the study was completed. Participants were recruited via newspaper ads or via the endocrinological outpatient clinic. Inclusion criteria comprised a BMI > 27 kg/m^2^ and postmenopausal status. The presence of concomitant untreated medical, neurological, and psychiatric diseases which may interfere with the planned interventions, such as instable coronary heart disease, kidney and liver disease, systemic infections, myopathy, food allergies, endocrinological disorders, and hypertension (systolic blood pressure > 180 mm Hg, diastolic blood pressure > 110 mm Hg) were excluded by medical history and physical examination. Lab screening was performed to rule out abnormal thyroid function and hypercortisolism using 1 mg dexamethasone suppression test. Individuals with recent weight changes of more than five kg during the last two months, with changes of smoking habits or diet behavior during the last three months were excluded. Participants with synthetic thyroid medications were not excluded as long as they were clinically euthyroid. The post-menopause status was ensured via medical history (cessation of the monthly period) and follicle stimulating hormone (FSH) screens if the status was unclear.

### Study design

This outpatient study was conducted in the Metabolic Research Unit of the Charité Universitätsmedizin Berlin. The flow chart of the trial is shown in Figure 1. Subjects were randomized into an intervention or a control group. We used a stratified randomization scheme based on 3 BMI (body mass index: weight [kg] / height [m]^2^) strata at baseline. Recruiting study nurses and physicians were blinded for the allocation sequence until inclusion of the participant. The allocation sequence was made via a random computer-generated list by the study PI and the statistician. Recruitment and allocation to the interventions was done by the study physicians.

**Figure 1.**
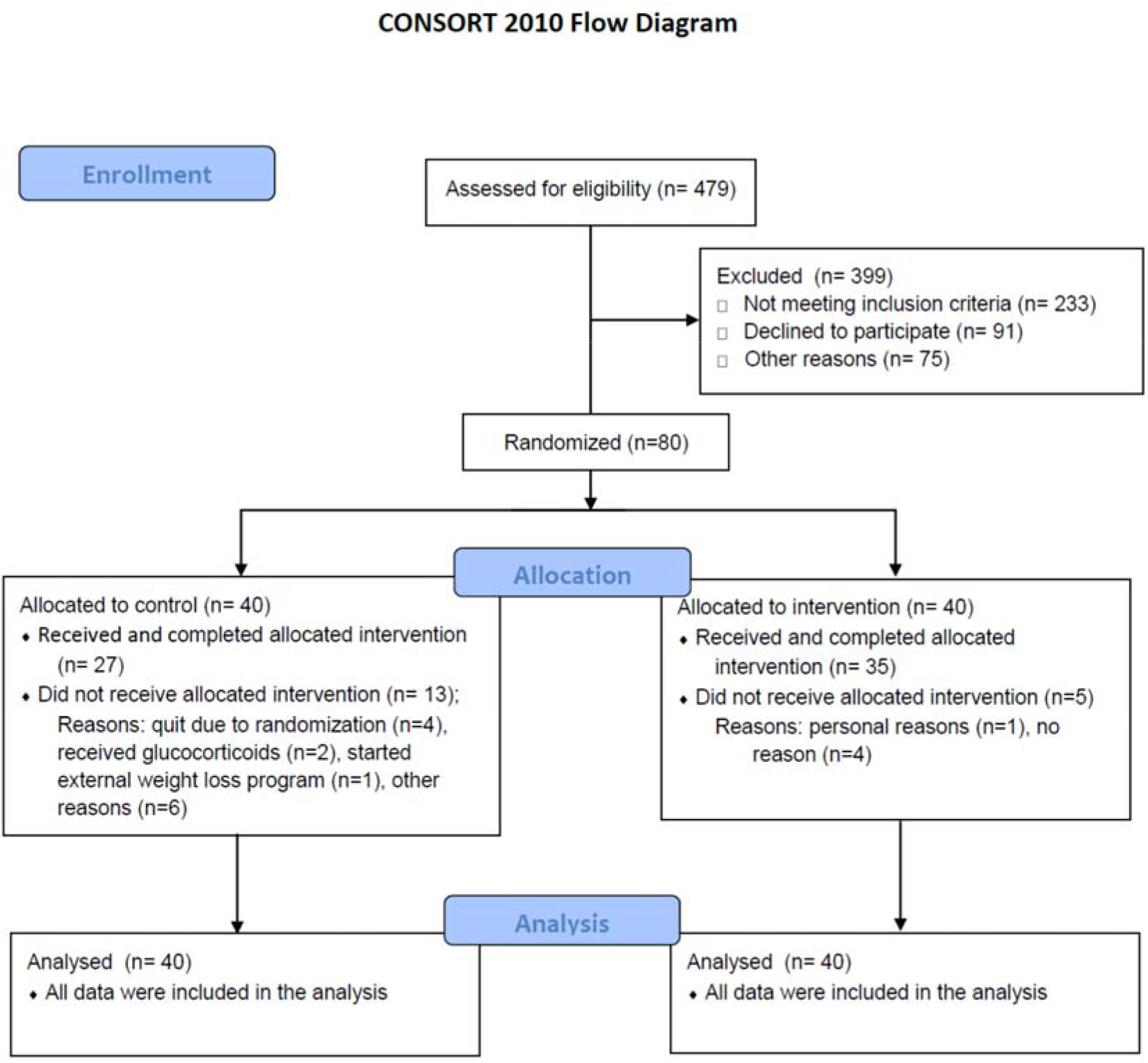
Flow chart of the randomized controlled trial.

After baseline phenotyping (specified below), participants belonging to the intervention group underwent a medically supervised weight loss and lifestyle intervention program. In detail, they were placed on an 800 kcal/d liquid diet (Optifast 2®, Nestlé HealthCare Nutrition GmbH, Frankfurt/Main, Germany) accompanied by weekly group meetings and weight loss counselling for 2 months. Participants were advised to consume the formula diet exclusively (daily consumption of five packets with 160 kcal each: 20 g carbohydrates, 14 g proteins, and 3 g fat dissolved in 300 ml water) without consuming any additional food. The eight weeks of formula diet were followed by four weeks in which the formula diet was substituted by a calorie reduced healthy diet to facilitate further weight loss. During this period participants were instructed to consume a balanced mix of macronutrients in accordance to the guidelines of the German Society for Nutrition (50-55% carbohydrates, 15-20 % proteins, and 30 % fat). The recommended daily calorie intake during these four weeks was based on the measured resting energy expenditure. Weekly counseling meetings were continued. Additionally, participants were encouraged to increase physical activity to 150 min of activity per week at least and step counting devices were handed out to monitor and motivate for physical activity. They were instructed to take at least 10,000 steps per day. However, no specific intervention regarding physical exercise was performed.

At the end of the weight loss period of 12 weeks (M3), phenotyping was repeated. Subsequently, participants were instructed to maintain their weight by changing daily caloric intake goals to a total calorie intake of the current resting metabolic rate plus 500 kcal. Adjustments of intended individual energy intake were made according to body weight development to achieve weight stabilization for another 4 weeks. After this weight stabilization phase (M4), initial phenotyping was repeated again.

Volunteers of the control group were instructed to keep their weight stable during the entire period of 4 months, which was ensured during monthly visits and nutritional counselling, which aimed for weight stabilization (resting metabolic rate plus 500 kcal). Phenotyping was performed in analogy to the intervention group at baseline (M0), after 3 months (M3) and after 4 months (M4). Further details of the intervention are provided in the supplements.

#### Follow up period

The 4-month randomized intervention period was followed by a free living period without any further active intervention in all subjects. Reevaluation including body weight and body composition was performed at month 12 (M12) and month 24 (M24).

### Phenotyping

A comprehensive phenotyping was performed at M0, M3 and M4. During each phenotyping time point, fasting blood was collected between 08.00 and 09.00 am after overnight fasting. Blood samples were centrifuged, and plasma and serum samples were frozen immediately at −80°C.

Each subject participated in a five-day protocol (M0-M4). This included anthropometric, hormonal and metabolic evaluation. All phenotyping procedures were performed after a 10-12 h overnight fast.

Glucose regulation was assessed by a 75g oral glucose challenge and subsequent blood sampling at 0, 30, 60, 90, 120 and 180 min. Homeostasis model assessment insulin resistance index (HOMA-IR) was used for approximation of insulin resistance^17^. Waist circumference was measured using a non-elastic tape measure three times consecutively and the mean was used for consecutive analyses. Body composition including fat mass (FM) and lean mass (LBM) was assessed by air displacement plethysmography (BodPod, COSMED Deutschland GmbH, Fridolfing, Germany). We further assessed 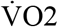 consumption and 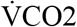 production by indirect calorimetry to calculate changes in energy expenditure (EE), resting metabolic rate (RER), and carbohydrate and fat oxidation rates. For measuring these parameters at rest, we used a canopy system (Quark RMR, COSMED Deutschland GmbH, Fridolfing, Germany). Diet induced thermogenesis (DIT) was assessed in a metabolic chamber. This chamber is a comfortable, airtight room (width: 2.5 m, depth: 2.0 m, height: 2.2 m, 11 m^3^) that is constantly supplied with fresh air like an open-circuit indirect calorimeter. In brief, 10h fasted volunteers entered the metabolic chamber at approximately 8 am and after a calibration phase of roughly 2 h, O_2_ consumption and CO_2_ production rates were measured continuously over a 45 min interval while patients were seated in a comfortable chair. Afterwards a mixed meal test was performed. Energy content of the mixed meal (Optifast 2®, Nestlé HealthCare Nutrition GmbH, Frankfurt/Main, Germany) reflected 33% of the individual resting energy expenditure (REE) of each subject at this time point. Postprandial thermogenesis was calculated up to 180 minutes after meal intake as previously described^18^, In brief, the difference between postprandial energy expenditure and resting energy expenditure was used to assess the thermic effect of the meal. DIT was calculated by dividing the thermic effect of the meal (calories expended above REE after ingestion of the mixed meal) by energy content of the individual mixed meal.

In addition, hyperinsulinemic euglycemic clamps were performed at a separate day at M0, M3 and M4 as described previously^19,20^. In brief, 40 mIU· m^-2^· min^-1^ human insulin (Actrapid®, Novo Nordisk, Bagsvaard, Denmark) and a variable infusion of 10 % glucose (Serag Wiessner, Naila, Germany) was used. Capillary glucose concentration was monitored every 5 minutes and was maintained between 4.0 and 4.9 mmol/l via variation of the glucose infusion rate. Blood samples were collected before the clamp and at least two hours after starting the clamp during steady-state conditions.

Total daily energy intake (TEI) was assessed by four-day food records at M0, M3 and M4. Therefore, volunteers were instructed to keep a food diary on every food item (including the exact weight or an equivalent estimation) that was consumed throughout these days. Nutrient content was analysed using Prodi v6.2 Expert software. The Souci-Fachmann-Kraut-Database 2005 and the German Federal Nutrition Key (Bundeslebensmittelschlüssel) Version 3.01 were used as nutrient databases. As these databases do not provide nutrient data for every kind of food item, it was necessary in some cases to select a similar food instead of the actual food named in the dietary record. These analyses were performed by one individual to ensure the best possible standardization.

### Laboratory tests

Laboratory analyses for routine clinical parameters were performed using established methods. Capillary blood glucose was measured using the glucose oxidase method (Dr. Müller Super GL, Freital, Germany). Serum insulin was measured using enzyme-linked immunosorbent assays (ELISA; Mercodia, Uppsala, Sweden) (intra-assay coefficient of variability (CV) 2.8 −4.0%, inter-assay CV 4 −5%). HbA1c was measured using a DCA 2000 Analyzer (Siemens) in our research unit. Leptin was analyzed using commercial ELISA (R&D Systems, Abingdon, UK).

### Sample size

Previous studies in adults report that a weight reduction by a change in lifestyle over 3 months is accompanied by a reduction in muscle mass from 3.2 to 6.9 %^21,22^. Assuming an effect of at least 3.2% and a variance of 3.4%, the power calculation yields a case number of 19 subjects per treatment arm (significance level 0.05 two-sided, power 80%, nquery 7.0). This effect difference refers to the end of the study intervention (maintenance phase 4 weeks after the end of the weight reduction program, M4). With an expected drop out number of 25 % during the weight loss phase and 15 % in the 4-week maintenance period, respectively, a total of at least 30 adults per treatment group should be included in the trial at the time of randomization.

### Outcomes

The co-primary endpoints were changes of myocellular insulin sensitivity assessed by euglycemic hyperinsulinemic clamp and changes of lean body mass assessed by air displacement plethysmography at time points M3 and M4 compared to M0. This randomized trial was explicitly performed to generate and evaluate novel hypotheses about the effects and mechanisms of a weight reduction. Therefore a number of secondary endpoints were defined which will be investigated in subsequent studies.

### Statistical analysis

We calculated body mass index (BMI) as the ratio between weight in kilograms and the square of the height in meters. Resting energy expenditure (REE) per LBM (REE_LBM_) was calculated by dividing REE (Kcal/d) by LBM (kg). Similar, leptin per FM (leptin_FM_) was calculated by dividing leptin (pg/ml) by FM (kg).

Skeletal muscle insulin sensitivity was assessed by dividing the average glucose infusion rate (GIR, mg glucose/min) during the steady state of the hyperinsulinemic euglycemic clamp by body weight (M-value). The insulin sensitivity index (ISI_Clamp_) was calculated as ratio of glucose metabolized during the steady-state period (M-value) to mean serum insulin concentration (I, mU/l) in this period of the euglycemic clamp. Whole body insulin sensitivity was estimated by HOMA-IR, which was calculated as previously described^17^. Changes of BMI, FM, LBM, HOMA-IR, and ISI_Clamp_, REE, REE_LBM_, DIT and TEI between each phenotyping time point were expressed as absolute values and labelled as ΔBMI, ΔFM, ΔLBM, ΔHOMA-IR, ΔISI_Clamp_, ΔREE, ΔREE_LBM_, ΔDIT and ΔTEI.

Statistical procedures were performed using SPSS version 25.0 (SPSS Inc., Chicago, IL, USA), and R-software package (version 4.0.3). Paired Student’s t-test for normally distributed data and Wilcoxon test for skewed data were used for within-group comparison between M4 and M24. Between-group comparisons at baseline were made via Wilcoxon-rank sum test. These data are presented as median and limits of the interquartile range (IQR: 25th – 75th percentile) and plotted as raw values unless stated otherwise. Data of the mixed-effect models are presented as mean and 95%CI. Results were considered significant, if the two-sided α was below 0.05.

Time courses of all variables between M0 and M4 were analyzed using a mixed-model, repeated-measures analyses of variance using the R package lmerTest (version 3.1-3). For each outcome variable, a separate mixed-model was created. The outcomes were adjusted to age and the interaction between squared time and group. We included participants as a random intercept term to account for the correlation of repeated measures within individuals. The here reported data are based on per protocol analysis including data of all available participants at the corresponding time point. Analysis of primary outcome was additionally performed as intention-to treat analysis with the use of the multiple imputation technique for missing data with the R software package mice (version 3.12.0). We created 10 imputed data sets with a maximum of 30 iterations. All imputations were done using the default predictive mean matching method from mice, despite of age, which was imputed using the last observation carried forward method. The results of the mixed model analysis with the imputed datasets are shown in the supplement.

Multivariate linear regression models were performed to analyze the impact of relative changes of ISI_Clamp_, LBM, REE, REE_LBM_ and DIT between M0 and M3 (ΔISI_Clamp-M0M3_, ΔLBM_M0M3_, ΔREE_M0M3_, ΔREE_LBM-M0M3_ and ΔDIT_M0M3_) or between M3 and M4 (ΔISI_Clamp-M3M4_, ΔLBM_M3M4_, ΔREE_M3M4_, ΔREE_LBM-M3M4_ and ΔDIT_M3M4_) or the corresponding levels at M3 or M4 on relative regain of FM between M4 and M24 (ΔFM_M4M24_). These models also included age and FM at M4 (FM_M4_) as potential confounders.

### Study approval

The study protocol was approved by the Institutional Review Board of the Charité Medical School and all subjects gave written informed consent. The trial was registered at www.clinicaltrials.gov under NCT01105143.

## 3. Results

80 overweight and obese postmenopausal women were enrolled in this trial. 35 of 40 participants randomized in the intervention group completed the allocated intervention whereas 27 of the 40 controls completed the treatment period. The overall retention rate was 77.5 %. A flow chart of the trial is shown in figure 1.

At baseline, subjects had an age of 59.0 (54.0 −64.0) years. Overweight was seen in 10 % of the randomized subjects, while 50%, 31% and 9% were characterized by obesity class I, II and III, respectively. The median BMI was 33.8 (32.2 −36.9) kg/m^2^ and relative fat mass was 48.7 (46.4 −52.3) %. Estimates of obesity, body composition, lipid metabolism, insulin resistance, liver function as well as energy and substrate utilization did not differ between both groups. Baseline characteristics of both groups are shown in table 1.

**Table 1:** Baseline characteristics of participants. Metabolic and anthropometric parameters of the randomized participants at baseline. Results are presented as median and interquartile range (IQR).

| Parameter | Intervention group |  |  | Control group |  | p value* |
| --- | --- | --- | --- | --- | --- | --- |
|  | median | (IQR) |  | median | (IQR) |  |
| Participants [n] | 40 |  |  | 40 |  |  |
| Age [yr] | 57.0 | (54.0-62.0) |  | 61.5 | (54.3-64.0) | n.s. |
| BMI [kg/m<sup>2</sup>] | 33.5 | (32.2-36.7) |  | 34.1 | (32.3-37.1) | n.s. |
| Fat mass [kg] | 44.4 | (41.8-48.5) |  | 46.2 | (41.4-50.4) | n.s. |
| Fat mass [%] | 49.4 | (46.1-52.6) |  | 47.8 | (46.5-51.6) | n.s. |
| Lean mass [kg] | 46.9 | (42.9-50.0) |  | 48.1 | (44.7-51.3) | n.s. |
| Lean mass [%] | 50.7 | (47.4-53.9) |  | 52.2 | (48.4-53.5) | n.s. |
| Waist circumference [cm] | 104.0 | (99.0-111.0) |  | 106.0 | (99.0-117.0) | n.s. |
| Hip circumference [cm] | 117.0 | (111.0-123.0) |  | 117.0 | (113.0-123.0) | n.s. |
| waist-hip-ratio | 0.89 | (0.84-0.92) |  | 0.91 | (0.87-0.96) | n.s. |
| Systolic blood pressure [mmHg] | 125.3 | (116.3-147.3) |  | 128.8 | (118.0-140.9) | n.s. |
| Diastolic blood pressure (mmHg) | 68.0 | (62.0-73.7) |  | 70.0 | (64.3-73.9) | n.s. |
| HbA1c [%] | 5.4 | (5.2-5.8) |  | 5.6 | (5.3-5.9) | n.s. |
| HOMA-IR<sup>#</sup> | 2.15 | (1.49-4.82) |  | 2.64 | (1.61-4.01) | n.s. |
| ISI<sub>Clamp</sub> [mg•kg<sup>-1</sup>•min<sup>-1</sup>/(mU•l<sup>-1</sup>)] | 0.062 | (0.034-0.079) |  | 0.048 | (0.031-0.067) | n.s. |
| Fasting glucose [mg/dl] | 99.6 | (94.3-108.0) |  | 103.5 | (94.7-117.0) | n.s. |
| Total cholesterol [mmol/l] | 5.2 | (4.6-6.0) |  | 5.3 | (4.9-6.5) | n.s. |
| HDL cholesterol [mmol/l] | 1.5 | (1.2-1.6) |  | 1.3 | (1.2-1.5) | n.s. |
| LDL cholesterol [mmol/l] | 3.1 | (2.7-3.5) |  | 3.2 | (2.8-3.9) | n.s. |
| Triacylglycerol [mmol/l] | 1.2 | (0.9-1.7) |  | 1.4 | (1.0-1.9) | n.s. |
| Alanine transaminase [U/l] | 16.4 | (12.4-25.1) |  | 18.5 | (13.6-24.9) | n.s. |
| Aspartate transaminase [U/l] | 22.3 | (17.8-27.2) |  | 23.3 | (19.7-28.3) | n.s. |
| Leptin [ng•ml<sup>-1</sup>] | 53.3 | (36.6-71.6) |  | 44.2 | (33.9-60.1) | n.s. |
| Leptin per fat mass [ng•ml<sup>-1</sup>•kg<sup>-1</sup>] | 1.13 | (0.84-1.55) |  | 1.03 | (0.78-1.30) | n.s. |
| Total energy intake [kcal•d<sup>-1</sup>] | 1937 | (1662-2767) |  | 2102 | (1788-2522) | n.s. |
| Resting energy expenditure [kcal•d<sup>-1</sup>] | 1487 | (1392-1629) |  | 1585 | (1454-1762) | n.s. |
| Respiratory coefficient | 0.76 | (0.73-0.79) |  | 0.76 | (0.74-0.80) | n.s. |
| Diet induced thermogenesis [%] | 12.0 | (9.3-14.5) |  | 10.5 | (8.0-12.5) | n.s. |

\*Wilcoxon rank-sum test; <sup>#</sup>Homeostatic Model Assessment inferring insulin resistance.

During the weight loss intervention, there was a substantial reduction in TEI within the intervention group between M0 and M3, while the TEI remained stable between M3 and M4. In contrast, energy intake was not changed within the control group throughout the study. As intended, BMI declined in the intervention group between M0 and M3, while no change was seen in the control group (Table 2). The between group comparison revealed a stronger BMI reduction between M0 and M3 as well as lower BMI levels at M3 in the intervention group (Table 3). There was no change of BMI in both groups beyond the weight loss period resulting in a comparable between-group difference at M3 and at M4. These changes were accompanied by a comparable course of FM, even if a clinical not important but statistical significant FM reduction was also seen between M0 and M3 in controls. The primary outcome LBM was reduced between M0 and M3 in the intervention group and remained unchanged between M3 and M4. No modification of LBM was seen within control subjects throughout the course of the randomized treatment period.

**Table 2:**
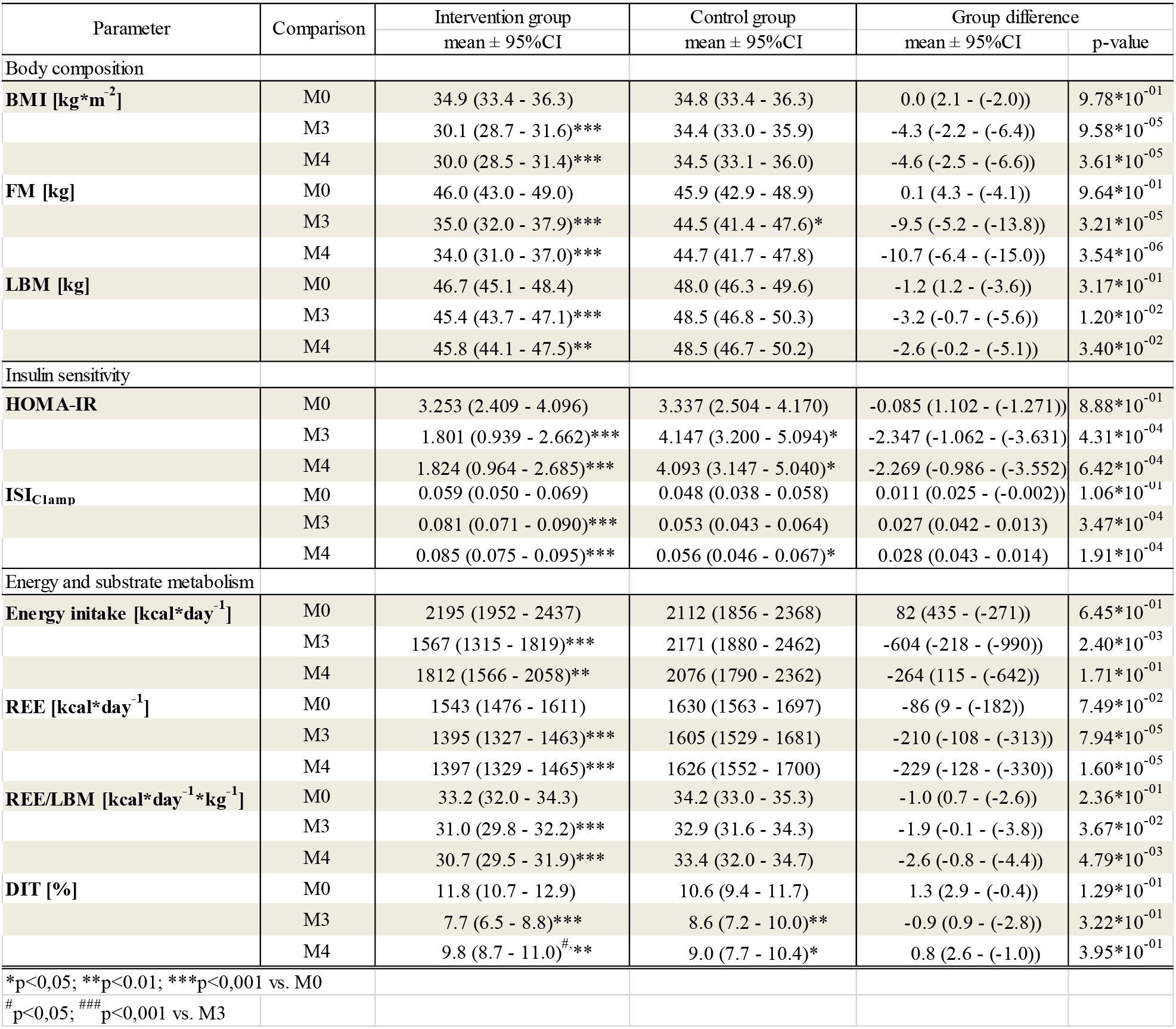
Estimates of body composition, insulin sensitivity and energy metabolism (mean and 95% CI) and estimated mean differences within per-protocol analysis. Intervention effects reported as estimated marginal means and estimated mean differences (intervention minus control) based on mixed-model, repeated-measures analysis of variance adjusted for treatment group and age.

| Parameter | Comparison | Intervention group | Control group | Group difference |  |
| --- | --- | --- | --- | --- | --- |
|  |  | mean ± 95%CI | mean ± 95%CI | mean ± 95%CI | p-value |
| Body composition |  |  |  |  |  |
| BMI [kg*m<sup>-2</sup>] | M0 | 34.9 (33.4 - 36.3) | 34.8 (33.4 - 36.3) | 0.0 (2.1 - (-2.0)) | 9.78*10<sup>-01</sup> |
|  | M3 | 30.1 (28.7 - 31.6)*** | 34.4 (33.0 - 35.9) | -4.3 (-2.2 - (-6.4)) | 9.58*10<sup>-05</sup> |
|  | M4 | 30.0 (28.5 - 31.4)*** | 34.5 (33.1 - 36.0) | -4.6 (-2.5 - (-6.6)) | 3.61*10<sup>-05</sup> |
| FM [kg] | M0 | 46.0 (43.0 - 49.0) | 45.9 (42.9 - 48.9) | 0.1 (4.3 - (-4.1)) | 9.64*10<sup>-01</sup> |
|  | M3 | 35.0 (32.0 - 37.9)*** | 44.5 (41.4 - 47.6)* | -9.5 (-5.2 - (-13.8)) | 3.21*10<sup>-05</sup> |
|  | M4 | 34.0 (31.0 - 37.0)*** | 44.7 (41.7 - 47.8) | -10.7 (-6.4 - (-15.0)) | 3.54*10<sup>-06</sup> |
| LBM [kg] | M0 | 46.7 (45.1 - 48.4) | 48.0 (46.3 - 49.6) | -1.2 (1.2 - (-3.6)) | 3.17*10<sup>-01</sup> |
|  | M3 | 45.4 (43.7 - 47.1)*** | 48.5 (46.8 - 50.3) | -3.2 (-0.7 - (-5.6)) | 1.20*10<sup>-02</sup> |
|  | M4 | 45.8 (44.1 - 47.5)** | 48.5 (46.7 - 50.2) | -2.6 (-0.2 - (-5.1)) | 3.40*10<sup>-02</sup> |
| Insulin sensitivity |  |  |  |  |  |
| HOMA-IR | M0 | 3.253 (2.409 - 4.096) | 3.337 (2.504 - 4.170) | -0.085 (1.102 - (-1.271)) | 8.88*10<sup>-01</sup> |
|  | M3 | 1.801 (0.939 - 2.662)*** | 4.147 (3.200 - 5.094)* | -2.347 (-1.062 - (-3.631)) | 4.31*10<sup>-04</sup> |
|  | M4 | 1.824 (0.964 - 2.685)*** | 4.093 (3.147 - 5.040)* | -2.269 (-0.986 - (-3.552)) | 6.42*10<sup>-04</sup> |
| ISI<sub>Clamp</sub> | M0 | 0.059 (0.050 - 0.069) | 0.048 (0.038 - 0.058) | 0.011 (0.025 - (-0.002)) | 1.06*10<sup>-01</sup> |
|  | M3 | 0.081 (0.071 - 0.090)*** | 0.053 (0.043 - 0.064) | 0.027 (0.042 - 0.013) | 3.47*10<sup>-04</sup> |
|  | M4 | 0.085 (0.075 - 0.095)*** | 0.056 (0.046 - 0.067)* | 0.028 (0.043 - 0.014) | 1.91*10<sup>-04</sup> |
| Energy and substrate metabolism |  |  |  |  |  |
| Energy intake [kcal*day<sup>-1</sup>] | M0 | 2195 (1952 - 2437) | 2112 (1856 - 2368) | 82 (435 - (-271)) | 6.45*10<sup>-01</sup> |
|  | M3 | 1567 (1315 - 1819)*** | 2171 (1880 - 2462) | -604 (-218 - (-990)) | 2.40*10<sup>-03</sup> |
|  | M4 | 1812 (1566 - 2058)** | 2076 (1790 - 2362) | -264 (115 - (-642)) | 1.71*10<sup>-01</sup> |
| REE [kcal*day<sup>-1</sup>] | M0 | 1543 (1476 - 1611) | 1630 (1563 - 1697) | -86 (9 - (-182)) | 7.49*10<sup>-02</sup> |
|  | M3 | 1395 (1327 - 1463)*** | 1605 (1529 - 1681) | -210 (-108 - (-313)) | 7.94*10<sup>-05</sup> |
|  | M4 | 1397 (1329 - 1465)*** | 1626 (1552 - 1700) | -229 (-128 - (-330)) | 1.60*10<sup>-05</sup> |
| REE/LBM [kcal*day<sup>-1</sup>*kg<sup>-1</sup>] | M0 | 33.2 (32.0 - 34.3) | 34.2 (33.0 - 35.3) | -1.0 (0.7 - (-2.6)) | 2.36*10<sup>-01</sup> |
|  | M3 | 31.0 (29.8 - 32.2)*** | 32.9 (31.6 - 34.3) | -1.9 (-0.1 - (-3.8)) | 3.67*10<sup>-02</sup> |
|  | M4 | 30.7 (29.5 - 31.9)*** | 33.4 (32.0 - 34.7) | -2.6 (-0.8 - (-4.4)) | 4.79*10<sup>-03</sup> |
| DIT [%] | M0 | 11.8 (10.7 - 12.9) | 10.6 (9.4 - 11.7) | 1.3 (2.9 - (-0.4)) | 1.29*10<sup>-01</sup> |
|  | M3 | 7.7 (6.5 - 8.8)*** | 8.6 (7.2 - 10.0)** | -0.9 (0.9 - (-2.8)) | 3.22*10<sup>-01</sup> |
|  | M4 | 9.8 (8.7 - 11.0)<sup>#</sup>** | 9.0 (7.7 - 10.4)* | 0.8 (2.6 - (-1.0)) | 3.95*10<sup>-01</sup> |
| *p<0,05; **p<0.01; ***p<0,001 vs. M0 |  |  |  |  |  |
| <sup>#</sup> p<0,05; <sup>###</sup> p<0,001 vs. M3 |  |  |  |  |  |

**Table 3:** Change of body composition, insulin sensitivity and energy metabolism (mean and 95% CI) within per-protocol analysis. Changes were reported as estimated mean differences based on mixed-model, repeated-measures analysis of variance adjusted for treatment group and age.

| Parameter | Comparison | Intervention group | Control group | Group difference |  |
| --- | --- | --- | --- | --- | --- |
|  |  | mean ± 95%CI | mean ± 95%CI | mean ± 95%CI | p-value |
| Body composition |  |  |  |  |  |
| ΔBMI | M0-M3 | -4.7 (-5.1 - (-4.3)) | -0.4 (-0.8 - 0.0) | -4.3 (-4.9 - (-3.7)) | 8.19*10<sup>-29</sup> |
|  | M3-M4 | -0.2 (-0.6 - 0.2) | 0.1 (-0.4 - 0.5) | -0.3 (-0.9 - 0.3) | 0.355 |
| ΔFM [kg] | M0-M3 | -11.0 (-12.1 - (-10.0)) | -1.4 (-2.8 - (-0.1)) | -9.6 (-11.3 - (-7.9)) | 1.24*10<sup>-19</sup> |
|  | M3-M4 | -0.9 (-2.1 - 0.2) | 0.3 (-1.1 - 1.6) | -1.2 (-3.0 - 0.6) | 0.184 |
| ΔLBM [kg] | M0-M3 | -1.4 (-2.0 - (-0.8)) | 0.6 (-0.2 - 1.3) | -1.9 (-2.9 - (-1.0)) | 6.44*10<sup>-05</sup> |
|  | M3-M4 | 0.5 (-0.1 - 1.1) | -0.0 (-0.8 - 0.7) | 0.5 (-0.4 - 1.5) | 0.285 |
| Insulin sensitivity |  |  |  |  |  |
| ΔHOMA-IR | M0-M3 | -1.452 (-2.090 - (-0.814)) | 0.810 (0.057 - 1.563) | -2.262 (-3.246 - (-1.278)) | 1.28*10<sup>-05</sup> |
|  | M3-M4 | 0.024 (-0.623 - 0.670) | -0.054 (-0.844 - 0.736) | 0.077 (-0.943 - 1.098) | 0.881 |
| ΔISI<sub>Clamp</sub> | M0-M3 | 0.021 (0.014 - 0.028) | 0.005 (-0.003 - 0.013) | 0.016 (0.005 - 0.026) | 3.40*10<sup>-03</sup> |
|  | M3-M4 | 0.004 (-0.003 - 0.011) | 0.003 (-0.005 - 0.011) | 0.001 (-0.010 - 0.012) | 0.843 |
| Energy and substrate metabolism |  |  |  |  |  |
| ΔEnergy intake [kcal*day<sup>-1</sup>] | M0-M3 | -628 (-895 - (-360)) | 59 (-244 - 361) | -686 (-1089 - (-283)) | 1.03*10<sup>-03</sup> |
|  | M3-M4 | 245 (-18 - 509) | -95 (-404 - 214) | 340 (-66 - 746) | 9.94*10<sup>-02</sup> |
| ΔREE [kcal*day<sup>-1</sup>] | M0-M3 | -149 (-202 - (-95)) | -25 (-89 - 40) | -124 (-207 - (-41)) | 3.85*10<sup>-03</sup> |
|  | M3-M4 | 2 (-51 - 55) | 21 (-43 - 85) | -19 (-102 - 64) | 0.654 |
| ΔREE/LBM [kcal*day<sup>-1</sup>*kg<sup>-1</sup>] | M0-M3 | -2.2 (-3.3 - (-1.0)) | -1.2 (-2.6 - 0.2) | -1.0 (-2.7 - 0.8) | 0.347 |
|  | M3-M4 | -0.3 (-1.4 - 0.9) | 0.4 (-1.0 - 1.8) | -0.7 (-2.5 - 1.1) | 0.446 |
| ΔDIT [%] | M0-M3 | -4.1 (-5.3 - (-3.0)) | -2.0 (-3.4 - (-0.5)) | -2.2 (-4.0 - (-0.3)) | 2.12*10<sup>-02</sup> |
|  | M3-M4 | 2.2 (1.0 - 3.4) | 0.5 (-1.0 - 1.9) | 1.7 (-0.2 - 3.6) | 7.84*10<sup>-02</sup> |

As reduction of FM was more pronounced than LBM loss, a presumably improved body composition could be detected in the intervention group after the weight loss period at M3. In detail relative LBM proportion increased by 6.3 (5.4 −7.3) % (p=1.59*10^−24^). Comparable changes of LBM were observed within the intention to treat analyses (Table S1 and S2).

As expected, subjects of the intervention group were characterized by a weight loss induced improvement of the second primary outcome ISI_Clamp_ at M3, which was not seen in the controls. Importantly, improved ISI_Clamp_ at M3 was similar at M4 indicating that improved muscle insulin sensitivity upon weight loss is mainly determined by changes in body composition and not energy balance status (Table 2 and 3). As expected, ISI_Clamp_ remained similar throughout all time points in the control group. Comparable changes of ISI_Clamp_ were observed within the intention to treat analyses (Table S1 and S2). The changes in ISI_Clamp_ in the intervention group were paralleled by a decline of HOMA-IR by 45% between M0 and M3, while a small increase of 24% was seen in control subjects.

Estimates of resting energy expenditure (REE and REE_LBM_) remained unchanged in the control group. In contrast, weight loss intervention induced an expected decline in REE which was completely preserved up to M4. This resulted in lower REE at M3 and M4 in the intervention group compared to control subjects. However, reduction of REE was stronger than expected by LBM reduction, as REE_LBM_ was also diminished by the weight loss intervention. Although this resulted in lower REE_LBM_ levels at M3 in the intervention group compared to control subjects, the between group difference of weight loss induced changes between M0 and M3 failed to reach significance. Weight maintenance without negative energy balance did not further modify REE_LBM_ (Table 3).

We next analyzed the effect of weight loss on DIT. We observed a decline of DIT in both groups between M0 and M3. This effect was more pronounced in the intervention group representing an effect of weight loss, changed body composition and negative energy balance at M3. Interestingly, this effect was partially reversed between M3 and M4 in the intervention group although body weight and body composition remained unchanged.

A substantial increase of both BMI and FM was detected between M4 and M24 within the intervention group (2.4 (0.7 – 3.9) kg/m^2^, p=4.4*10^−6^ and 6.3 (1.8 – 10.3) kg, p=2.8*10^−6^). In contrast neither BMI nor FM were changed between M4 and M24 in controls (−0.0 (−1.4 – 0.6) kg/m^2^, p=0.218 and −0.5 (−3.9 – 2.2) kg, p=0.258) (Figure 2).

**Figure 2.**
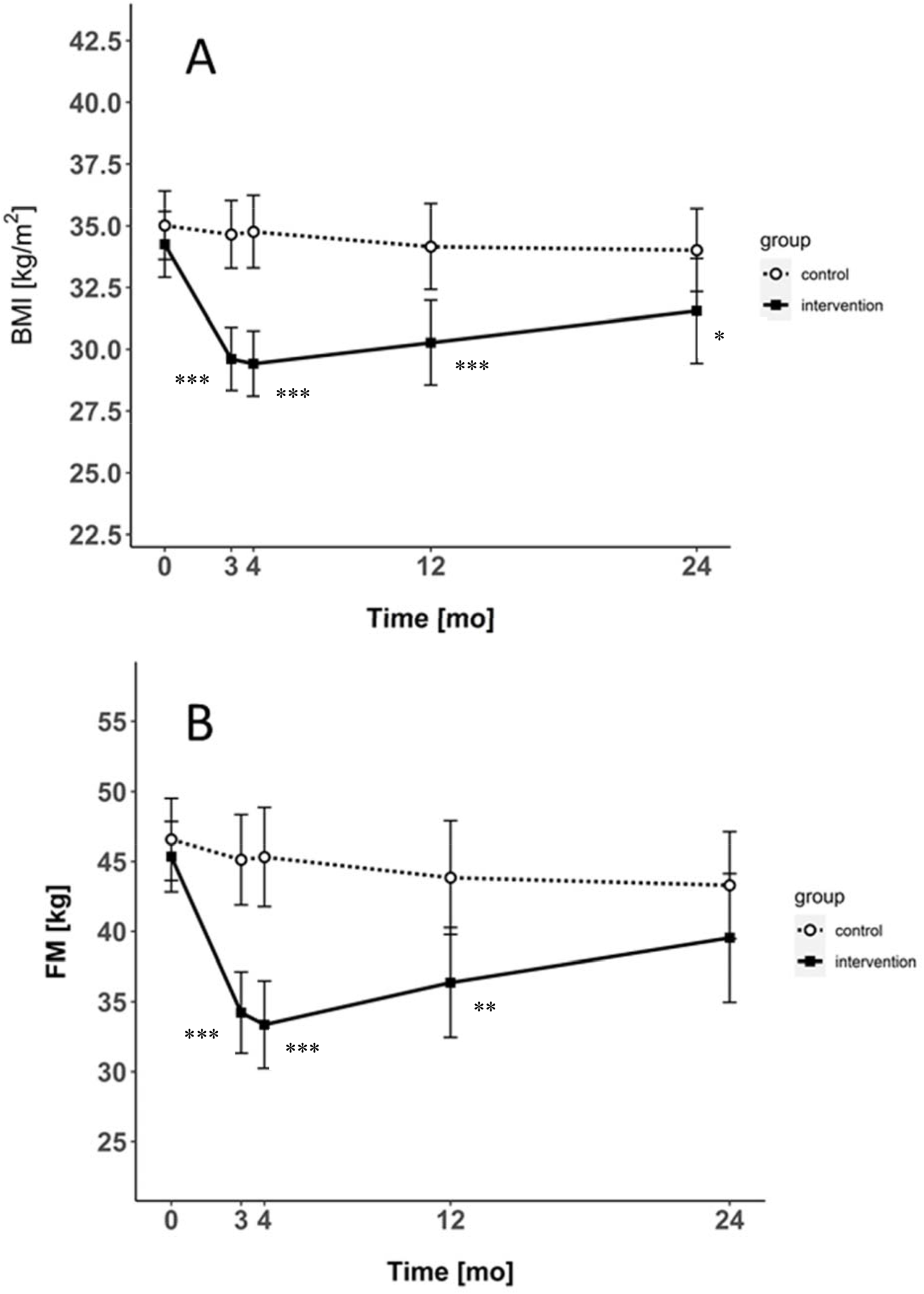
BMI (A) and FM (B) during the randomized controlled trial and follow-up. BMI (A) and FM (B) of the control (open circles) and intervention group (filled squares) during the randomized weight loss (M0 to M3), the maintenance period (M3 to M4) and the follow up (M4 to M24) analyzed per protocol. Raw values were plotted. Results are presented as mean ± 95% Confidence Intervals (CI). *p<0.05, **p<0.01 and ***p<0.001 control vs. intervention group based on Wilcoxon-rank sum test.

Finally, we aimed to separate effects induced by weight loss and concomitant negative energy balance on long-term development of obesity after weight loss. However, neither individual changes of the primary outcome parameters (LBM and ISI_Clamp_) between M0 and M3 and between M3 and M4 nor corresponding levels at M3 and M4 predicted long-term course of FM beyond M4 (ΔFM_M4M24_). Comparably, in the intervention group LBM, HOMA-IR and DIT at M3 and M4 as well as changes between the different time points were not predictive for ΔFM_M4M24_. However, lower REE and REE_LBM_ levels at M3 as well as the individual increase of REE and REE_LBM_ between M3 and M4 predicted a stronger ΔFM_M4M24_ (Table 3 and 4).

**Table 4:**
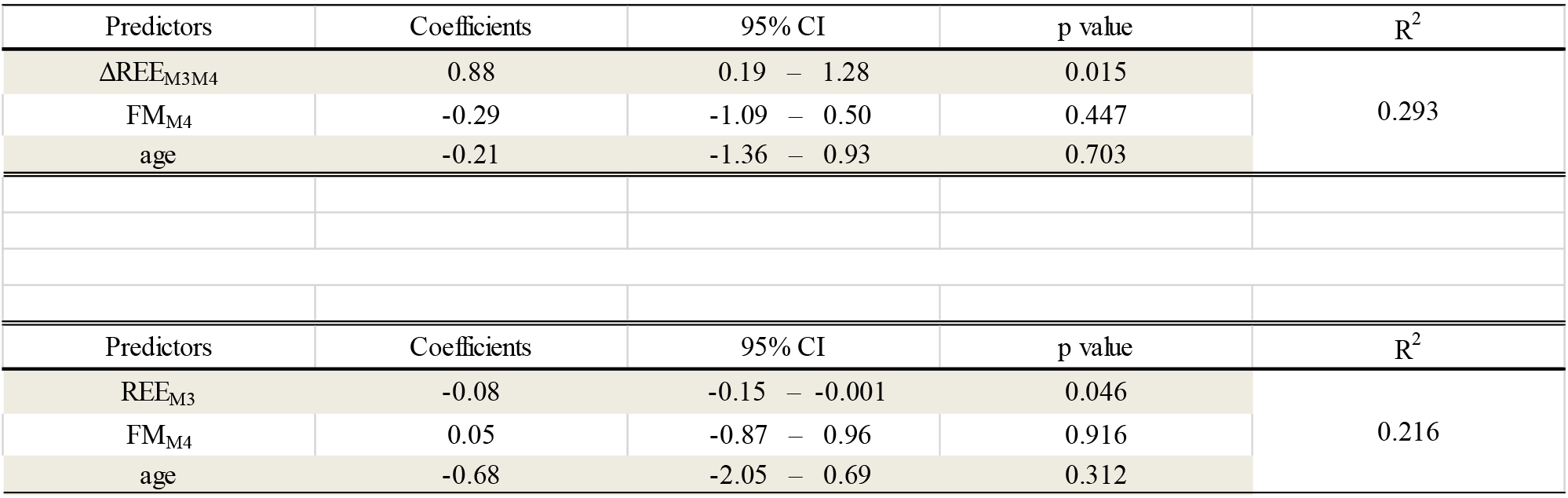
Impact of ΔREE_M3M4_ on ΔFM_M4M24_ (A) and REE_M3_ on ΔFM_M4M24_ (B).

| Predictors | Coefficients | 95% CI | p value | R<sup>2</sup> |
| --- | --- | --- | --- | --- |
| $\Delta\text{REE}_{\text{M3M4}}$ | 0.88 | 0.19 – 1.28 | 0.015 | 0.293 |
| $\text{FM}_{\text{M4}}$ | -0.29 | -1.09 – 0.50 | 0.447 | |
| age | -0.21 | -1.36 – 0.93 | 0.703 |  |
| Predictors | Coefficients | 95% CI | p value | R<sup>2</sup> |
| $\text{REE}_{\text{M3}}$ | -0.08 | -0.15 – -0.001 | 0.046 | 0.216 |
| $\text{FM}_{\text{M4}}$ | 0.05 | -0.87 – 0.96 | 0.916 | |
| age | -0.68 | -2.05 – 0.69 | 0.312 |  |

**Table 5:**
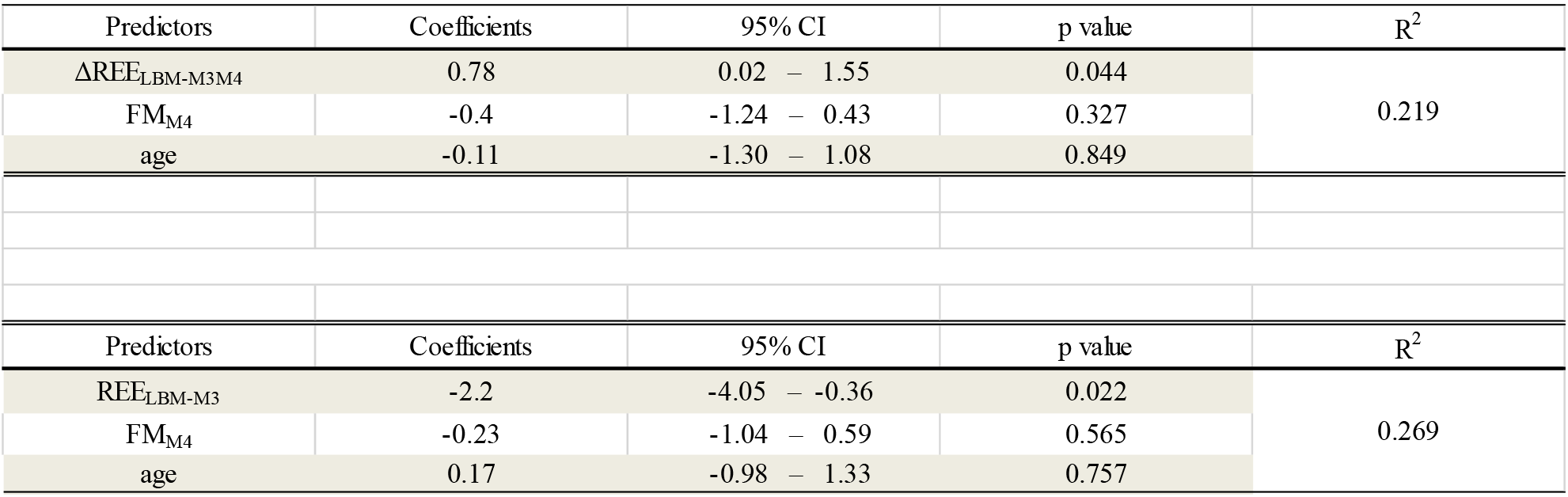
Impact of ΔREE_LBM-M3M4_ on ΔFM_M4M24_ (A) and REE_LBM-M3_ on ΔFM_M4M24_ (B).

## 4. Discussion

This randomized controlled clinical trial was initiated to explore mechanisms regulating body weight, energy balance, insulin sensitivity and body weight composition in a metabolically phenotyped cohort. We aimed to separate the effects of modified body weight and body composition versus the effect of a negative energy balance after weight loss. As expected, weight loss in the intervention group was accompanied by a reduction in fat mass and lean body mass. Notably both body weight and body composition did not change significantly during the stabilization period of four weeks, enabling us to dissociate the different effects of a negative energy balance during weight loss and a changed body composition after weight loss.

Skeletal muscle tissue plays a dominant role in insulin mediated glucose uptake and glucose metabolism. Although a decrease in myocellular lipid content is a previously described mechanism of weight loss induced improvement of insulin sensitivity, alterations of skeletal muscle mass during a weight reduction may also affect myocellular insulin sensitivity. In accordance with numerous dietary and bariatric weight loss strategies demonstrating an increase of insulin sensitivity^23,24^, we observed an improvement of insulin sensitivity after 3 months of weight loss. The unchanged insulin sensitivity after an additional month of preserved body weight and body composition indicates that the negative energy balance during weight loss has no modulating impact on insulin sensitivity. This is apparently different from the acute effect of 60h fasting, which resulted in a decline of myocellular insulin sensitivity, possibly induced by the fasting induced increase of FFA^25,26^. Beside neuroendocrine mechanisms regulating body weight, local aspects in fat and muscle are likely to play a role. Myocellular insulin sensitivity is known to regulate insulin mediated inhibition of lipolysis. However, modulation of insulin sensitivity did not predict future weight or fat mass regain. This indicates that other mechanisms in the process of adipose tissue metabolism may play a role, such as has been shown previously for atrial natriuretic peptide^1^ and sympathetic nervous system^10^.

As expected, and shown in numerous previous trials^27-31^, lean body mass and resting energy expenditure per metabolic mass was down-regulated after weight loss, a phenomenon not seen in the control group. This confirms previous findings from smaller non-randomized studies and thus supports the relevance of this adaptive mechanism. Negative energy balance itself apparently did not affect the downregulation of resting energy expenditure, as this downregulation persisted up to M4. In accordance with the downregulation of resting energy expenditure, we also observed a weight-loss-induced reduction of meal induced thermogenesis. Although this was also seen in controls, it was clearly more prominent in the weight loss group. However, anticipating a mean daily calorie intake of app. 1500-2000 kcal, the detected decline in DIT of 4.1 % would reflect a total decline in energy expenditure of 62-82 kcal/d. This might promote energy storage after food consumption and may represent a relevant mechanism counterbalancing weight loss and potentially drive body weight regain after successful weight loss. Comparable findings were also found in previous trials analyzing smaller groups after weight loss^28,29^. Although these trials did not report this difference as significant, it is likely the result of a smaller sample size and an overall moderate effect.

Even if estimates of resting energy expenditure were unchanged between M3 and M4, a considerable individual variability was detected. Comparable findings were already described for weight loss induced changes of energy expenditure^32^. We speculated that this variability might be relevant in the prediction of weight regain. In fact, a high difference of unadjusted and LBM adjusted resting energy expenditure between M3 and M4, which reflects the additional individual adaption of basal metabolism to the negative energy balance, was one of the predictors of long-term fat mass regain. Accordingly, low REE and REE_LBM_ directly measured after weight loss (i.e during negative energy balance), were also relevant predictors of fat mass regain. These data highlight the impact of metabolic adaption to negative energy balance, which is apparently more relevant for future weight regain than changes induced by weight loss itself. Our data support that some individuals develop a more thrifty and spendthrift phenotype during weight loss than others. In line with these findings, a well-designed but non-randomized small trial in 7 subjects indicated, that a higher decline in 24h energy expenditure during a 24h fasting period promotes a smaller weight loss during a subsequent dietary caloric restriction^33^. These diametrical phenotypes are respectively also characterized by a lower or stronger increase of 24h energy expenditure during overfeeding^34^.

The interpretation of our data is limited by some factors. Behavioral, social as well as environmental factors may have influenced the results of our trial^35-37^. Although we performed group sessions under standardized setting and the weight loss intervention based on standardized VLCD, we cannot exclude that these factors may have influenced our results due to the outpatient design of our trial. Given the known effect of different macronutrients on insulin sensitivity, this might have influenced our results as well, especially beyond the period of VLCD intake.

On the other hand, some strengths of the current trial should be mentioned. These include the large sample size, the randomized controlled design, the long duration of the intervention, subsequent observation and the comprehensive phenotyping including detailed repeated assessment of myocellular insulin sensitivity by hyperinsulinemic-euglycemic clamp, body composition by air displacement plethysmography and energy and substrate metabolism using a metabolic chamber. In particular, the comparison to a real control groups characterized by an almost unchanged body composition and metabolic phenotype throughout the trial improved the robustness of our analyses. In summary, our data demonstrated that improvement of insulin sensitivity and alteration of LBM during weight loss are mainly driven by changes in body weight and composition and not by energy balance status. Interestingly, our data indicate the existence of a thrifty phenotype during the period of negative energy balance, which is highly susceptible to future regain of fat mass after weight loss interventions. Characterization of underlying mechanism driving this variability may allow improvement and individualization of current weight loss strategies.

## Supporting information

Study protocol

## Data Availability

The datasets generated during and/or analyzed during the current study are available from the corresponding author on reasonable request.

## Author Contributions

L.S., J.S. and K.M. researched data and wrote the manuscript; L.S., U.Z., J.S. and K.M. were responsible for data analysis; J.B., M.B., R.JvS., N.S and S.D. researched data. J.S. and K.M. designed the study. All authors contributed to interpretation of the results. All authors critically read and edited drafts before submission. All authors read and approved the submitted version.

## Acknowledgments

We thank F. Schwerin, L. Franz, N. Huckauf and C. Kalischke for excellent technical assistance as well as Heike Berger for the support regarding nutritional counseling. We thank Nestlé HealthCare Nutrition GmbH, Frankfurt am Main, Germany for the opportunity to purchase the Optifast 2^®^ diet at a reduced price.

## Ethics approval and consent to participate

The study protocols were approved by the Institutional Review Board of the Charité Medical School (EA1/140/12) and all subjects gave written informed consent.

## Tables and Figures

**Table S1:** Estimates of LBM and ISI_Clamp_ (mean and 95% CI) and estimated mean differences within intention to treat analysis. Intervention effects reported as estimated marginal means and estimated mean differences (intervention minus control) based on mixed-model, repeated-measures analysis of variance adjusted for treatment group and age.

| Parameter | Comparison | Intervention group | Control group | Group difference |  |
| --- | --- | --- | --- | --- | --- |
| | | mean ± 95%CI | mean ± 95%CI | mean ± 95%CI | p-value |
| **LBM [kg]** | M0 | 46.9 (45.3 - 48.5) | 48.0 (46.3 - 49.6) | -1.1 (1.3 - (-3.4)) | $3.65 \cdot 10^{-01}$ |
| | M3 | 45.5 (43.9 - 47.2)*** | 48.4 (46.7 - 50.1) | -2.9 (-0.5 - (-5.3)) | $1.98 \cdot 10^{-02}$ |
| | M4 | 45.8 (44.2 - 47.5)** | 48.4 (46.7 - 50.0) | -2.5 (-0.2 - (-4.9)) | $3.30 \cdot 10^{-02}$ |
| **ISI<sub>Clamp</sub> [mg•kg<sup>-1</sup>•min<sup>-1</sup>/(mU•l<sup>-1</sup>)]** | M0 | 0.059 (0.050 - 0.069) | 0.047 (0.038 - 0.057) | 0.012 (0.026 - (-0.001)) | $7.64 \cdot 10^{-02}$ |
| | M3 | 0.079 (0.070 - 0.089)** | 0.054 (0.044 - 0.065) | 0.025 (0.040 - 0.010) | $1.26 \cdot 10^{-03}$ |
| | M4 | 0.085 (0.074 - 0.095)*** | 0.058 (0.047 - 0.069)* | 0.027 (0.042 - 0.012) | $6.26 \cdot 10^{-04}$ |
| *p<0,05; **p<0.01; ***p<0,001 vs. M0 |  |  |  |  |  |

**Table S2:**
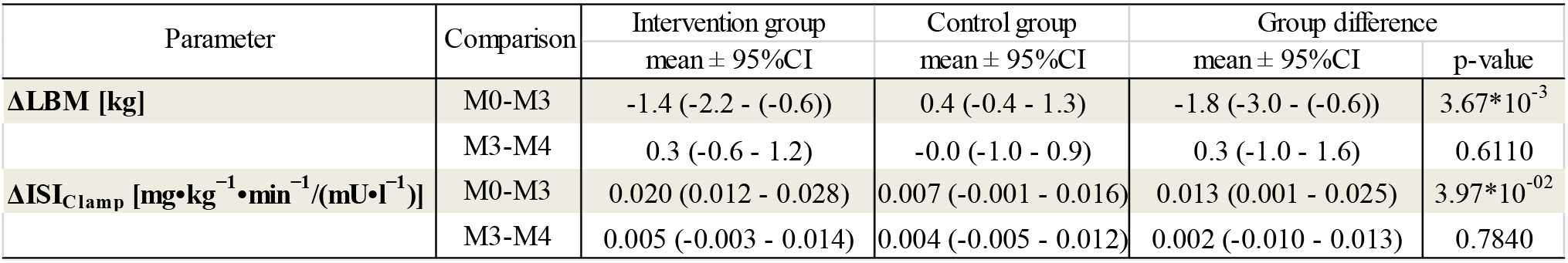
Change of LBM and ISI_Clamp_ (mean and 95% CI) within intention to treat analysis. Changes were reported as estimated mean differences based on mixed-model, repeated-measures analysis of variance adjusted for treatment group and age.

