## Supplementary material for "Thrifty energy phenotype predicts weight regain - results of a randomized controlled trial": Study protocol

**Analysis of hormonal and metabolic effects of a negative energy balance on the regulation of muscle mass and muscle function**

**"MMS (Muscle Metabolism Study)"**

**ClinicalTrials.gov Identifier: NCT01105143**

**1	Synopsis**

| **Title of the clinical study** | Analysis of hormonal and metabolic effects of a negative energy balance on the regulation of muscle mass and muscle function |
| --- | --- |
| **Type of the study** | Randomized controlled clinical trial |
| **Principal Investigator** | Prof. Dr. Knut Mai  Clinic of Endocrinology, Diabetes and Nutrition  Charité - Universitätsmedizin Berlin  Charitéplatz 1 , 10117 Berlin  Tel: +49/30/450-514252  |
| **Objective** | A randomized controlled weight loss intervention trial investigating the effects of acute and stabilzed weight loss on the hormonal regulation of skeletal muscle mass and function. A detailed and exploratory systems-based approach including characterization of energy metabolism, hormonal circuits, tissue histology, ex-vivo experimentation, immunophenotyping, DNA and RNA sequencing and characterization of the human gut microbiome will be applied to understand the complex physiology of the regulation of muscle mass and function during different states of energy balance. |
| **Trial endpoints** | **Primary endpoints**   1. Changes of myocellular insulin sensitivity (hyperinsulinemic clamp) during negative energy balance and during stabilized modification of body composition after weight loss. 2. Changes of skeletal muscle mass (air displacement plethysmography) during negative energy balance and during stabilized modification of body composition after weight loss.   **Secondary endpoints**   3. Effects on energy expenditure; Measurement of energy expenditure (kcal/d), postprandial thermogenesis (%) and respiratory coefficient (M0-M4) 4. Effects on myocellular and adipose tissue metabolism and substrate utilization; Measurement of myocellular and adipose metabolism using microdialysis (glycerol (µmol/l), lactate (mmol/l), pyruvate (µmol/l), glucose (mmol/l)) during oral glucose load (180 minutes) 5. Effects on myocellular and adipose tissue mRNA expression Analysis of myocellular and adipose mRNA expression (RNA sequencing) in counts 6. Weight regain; Analysis of body weight regain (BMI; kg/m^2^) during follow up 7. Fat mass; Analysis of body fat (kg and %) M0-M24 8. Assessment of the human gut microbiome before, during and after weight loss (negative energy balance) and during stabilized modification of body composition 4 weeks after weight loss via 16S rRNA sequencing and/or shotgun metagenomic pyrosequencing of the gut microbiota for composition and gene abundances 9. FFA during negative energy balance and during stabilized modification of body composition after weight loss; Measurement of fatty acids at baseline, during negative energy balance, during stabilized modification of body composition after weight loss and during follow up 10. Metanephrines during negative energy balance and during stabilized modification of body composition after weight loss; Measurement of metanephrines at baseline, during negative energy balance, during stabilized modification of body composition after weight loss and during follow up 11. Leptin during negative energy balance and during stabilized modification of body composition after weight loss; Measurement of leptin at baseline, during negative energy balance, during stabilized modification of body composition after weight loss and during follow up 12. Cortisol during negative energy balance and during stabilized modification of body composition after weight loss; Measurement of cortisol at baseline, during negative energy balance, during stabilized modification of body composition after weight loss and during follow up 13. Follistatin during negative energy balance and during stabilized modification of body composition after weight loss; Measurement of follistatin at baseline, during negative energy balance, during stabilized modification of body composition after weight loss and during follow up 14. Adiponectin during negative energy balance and during stabilized modification of body composition after weight loss; Measurement adiponectin at baseline, during negative energy balance, during stabilized modification of body composition after weight loss and during follow up 15. Natriuretic peptide during negative energy balance and during stabilized modification of body composition after weight loss; Measurement of natriuretic peptide at baseline, during negative energy balance, during stabilized modification of body composition after weight loss and during follow up. 16. Investigating associations between insulin sensitivity and immune cell composition during weight loss and genetic sequencing of the T-cell receptor 17. Comparison of metabolic parameters in different subcutaneous fat depots 18. IGF-1 during negative energy balance and during stabilized modification of body composition after weight loss; Measurement of IGF-1 at baseline, during negative energy balance, during stabilized modification of body composition after weight loss and during follow up 19. Analysis of predictive impact of several hormonal and metabolic parameters on body weight regain, course of insulin sensitivity and metabolism; The effect of measured parameters (see other endpoints) on long-term course of BMI, muscle mass, insulin sensitivity and energy expenditure will be analyzed using mathematical models |
| **Intervention** | Overweight or obese individuals will be randomised in two arms (intervention and control). The intervention group will undergo a life style intervention including 8 weeks on a 800 kcal per day formula diet followed by another 4 weeks of a calorie reduced normal diet accompanied by frequent nutritional counseling. After this period, individuals will be counseled to keep the weight stable for another 4 weeks. The control group will be advised to keep the initial weight stable during the whole study period. Follow-up includes a free-living period with no further instructions and study visits after 12 and 24 months. |
| **Inclusion Criteria** | Inclusion Criteria:   - BMI > 27 kg/m2 (adults) - postmenopausal state   Exclusion criteria:   - weight loss of more than 5kg in the last 2 months - unhealthy patients with: severe chronic diseases including cancer within the last 5 years, severe heart disease, severe impairment of hepatic or renal function, severe anaemia or disturbed coagulation - eating disorders or any other psychiatric condition that would interact with the trial intervention - malabsorption - acute or chronic infections - severe hypertension - myopathy - food allergies - any other uncontrolled endocrine disorder - changes of smoking habits, diets or medication that strongly affects energy homeostasis within the last 3 months prior to study inclusion |
| **Study duration** | 4 months intervention period and subsequently 20 months free-living follow-up period |
| **Statistical methods** | The primary and secondary outcomes will be analyzed by an intention to treat (ITT) analysis (primary analysis) as well as a per-protocol analysis (secondary analysis). The intention-to-treat (ITT) population is defined by all randomized patients who have participated in at least one study visit. Systems-based analyses will include multidimensional data processing and machine learning.  Prediction analyses will be performed using linear or logistic  regression models and/or machine learning algorithms. |
| **Number of subjects** | At least 80 female subjects |
| **Number of sites** | n=1 |
| **Data collection** | Data collection will be done by CRF |

**2	Summary**

Beside a loss in fat mass, weight reduction in most cases is characterized by substantial loss of muscle mass. The unintended reduction of muscle mass during weight reduction may be causally related with weight regain due to reduced energy requirements. Loss of muscle mass during weight loss is more pronounced than the regain of muscle mass during subsequent weight regain. This imbalance leads to the assumption, that a repetitive weight loss in combination with several periods of weight regaining (cycling) might be disadvantageous compared to a continuous state of obesity, since the cycling might result in decreased muscle mass compared to body fat. Thus, muscle tissue is crucially involved in short- and long-term weight regulation however the underlying mechanisms are yet not understood. We therefore conduct a randomized controlled clinical life style intervention trial to find critical factors that control muscle mass and function during weight loss. Eighty-one postmenopausal overweight or obese women will be recruited and randomized into two groups. Participants of the intervention group will undergo a weight reduction intervention (8 % expected weight loss) that will be implemented by lifestyle changes by means of increased physical activity and nutrition counseling. In addition, weight loss will be supported by placing study participants on an 800 kcal/d liquid diet during the first 8 weeks of intervention excluding any other food for this time period. Followed by 4 weeks of a calorie reduced normal diet, individuals will then be counseled to keep their weight stable for another 4 weeks, during this phase a negative energy balance is to be avoided. Nutritional counseling will take place once per week in the intervention group. The control group will be instructed to remain weight stable throughout the initial 4 months of the study and receive brochures with information about a healthy lifestyle. A further intervention will not take place in this group. Before the intervention (M0), after 3 months of weight loss (M3) and after the following 4 weeks of weight maintenance (M4), individuals will undergo metabolic phenotyping including measurements of body composition, glucose regulation and insulin sensitivity. Additionally, muscle and subcutaneous fat biopsies will be taken during each of these time points. Stool samples will be collected frequently throughout the study period to monitor changes within the human gut microbiota during the intervention. This phenotyping will be performed simultaneously in the control group. After 4 months of the intervention phase, all participants will enter the free-living ad libitum follow-up phase of the study without any further instructions regarding weight management and life style. Follow-up visits will take place after 12 months (M12) and 24 months (M24).

**3	Background and Rationale**

Obesity is a disease with increasing global importance. Both, the increasing availability of high-caloric foods and the shift in lifestyle to primarily sedentary activity with decreasing physical activity lead to a continuously increasing incidence among children and adults in our society. In the US, about 30 % of adults and 16 % of children are obese (BMI > 30 kg/m^2^) (1). According to the Federal Statistical Office, in Germany, about 35 % of the people currently suffer from overweight (BMI 25-30 kg/m^2^) and around 13 % from obesity. Obesity-related diseases are well known and include comorbidities such as type 2 diabetes mellitus, arterial hypertension and hyper- or dyslipidaemia, each of which often occur in patients with metabolic syndrome. Thus, an increased mortality was been reported for obese individuals.

Despite intensive efforts, both prevention and treatment of obesity remains difficult and is mostly inefficient. In this regard the impact of skeletal muscle on the regulation of bodyweight is poorly understood, although it has a pivotal role in the regulation of metabolism and most likely also for general health. Data indicate that loss of muscle mass is predictive of overall mortality (2). However a loss of muscle mass is not only a physiological consequence of aging, but can also be observed in obese individuals during intended weight reduction. While a correlation does exist between the reduction in fat mass and a partial loss of muscle mass during weight loss, the magnitude of this relation is rather low. It is currently consensus, that the unintended reduction of muscle mass during weight reduction is causally related with weight regain via reduced energy requirements. Remarkably, the loss of muscle mass during weight reduction is more pronounced than the regain of muscle mass during subsequent weight regain (3). This imbalance leads to the assumption, that a repetitive weight loss in combination with several periods of weight regaining (cycling) might be disadvantageous compared to a continuous state of obesity, since weight cycling might result in a decreased muscle to fat mass ratio over time.

The regulation and the metabolic significance of these processes are mostly unknown. As far as is known, a neuroendocrine network, including leptin, ghrelin, steroids and thyroid hormones, orchestraes processes that contribute to energy homeostasis and the regulation of fat mass. Such neuroendrocrine network may also be involved in muscle mass regulation. Thus, the knockout of myostatin, which is a circulating member of TGFβ-superfamily, leads to a profound increase of muscle mass in mice (4). Accordingly, overexpression of myostatin results in a wasting-syndrome, which is characterized by a significant loss of muscle mass (5). Moreover, other hormone systems may be involved, e.g. glucocorticoides since it has been shown that steroidmyopathie is associated with an elevated expression of myostatin (6). In addition, also growth hormones play a role in muscle mass regulation since growth hormone therapy in patients suffering from growth hormone deficiency leads to a reduction in body fat mass while muscle mass increases.

However, it is unclear which cells are primarily targeted by the hormonal signals. Some data point to satellite cells that might play a critical role for an increase in total muscle cell count. It has been shown that treatment with follistatin, which is an inhibitor of myostatin, leads to muscle hypertrophy via an activation of satellite cells (7). In addition, satellite cells can apparently differentiate into brown fat tissue under certain conditions (8,9) which indicates their potential critical role for the energy homeostasis.

Apart from lower energy expenditure as a result of muscle mass reduction, also other mechanisms within the skeletal muscle tissue might play a key role in the regulation of energy homeostasis. For instance, various cytokines, such as interleukin (IL)-6, IL-8, IL-15 and TNFα, are produced and secreted in the skeletal muscle tissue and can affect muscle mass as well as several metabolic processes (10). For instance, it has been shown for IL-6 and TNFα that the cytokine concentration correlates with the degree of obesity and insulin resistance (11,12). In addition, the release of IL-6 during physical activity enhances lipolysis in adipocytes (10). On the other hand, there are a number of adipocytokines, such as adiponectin, chemerin and leptin, which in turn can differently affect the skeletal muscle. Here, adiponectin was shown to improve muscle insulin sensitivity (13). Thus, various cytokines are regulated in obesity and weight loss, however, for most of them it is unclear how far and to what extent they might affect muscle mass.

Considering the fact that obesity has become one of the major global health problems, intervention strategies so far are usually only temporarily successful and the mechanisms that lead to regain of body weight after initial weight loss in the majority of patients are currently poorly understood, it seems imperative to investigate in detail the role of the skeletal muscle tissue in weight regulation.

Since active weight loss includes negative energy balance and changes in body composition, in this study, effects of weight loss during negative energy balance and effects of an altered body composition during neutral energy balance will be analyzed respectively.

Elucidation of the microbial and viral composition of human stool samples at given conditions are based on previous studies, which have demonstrated, that short-term changes of food intake or weight loss after negative energy balance affect the structure of the colonic microbiome (14,15). First data in humans show associations between gut bacteria and the effectiveness of nutrient absorption (14). Further studies in mice indicate that the colonic microbiome by itself has an impact on body composition (16). However, the regulation of gut microbial diversity is poorly understood. Beside nutrients as strong drivers of microbial diversity, bacteriophages (virus) might play a major role as well (17). So far, data addressing the characterization of the colonic virome in the context of a negative energy balance and weight loss are not available. The composition of the colonic microbiome before, during and after weight loss will provide essential information about dynamic relations between food, body composition and the intestinal microbiota.

In particular, with regard to associations between gut bacteria and nutrient absorption, it will be further investigated whether the composition of the gut microbiota is associated with long-term development of body weight.

The investigations of cognitive and morphological brain parameters in a subgroup of this study are based on epidemiological studies as well as on experimental animal models, both of which have shown that energy restriction decelerates the development of age-related functional deficits as well as the risk of disease for neurodegenerative disorders, such as M. Alzheimer and M. Parkinson. In animal models energy restricted diet results in lowered age-related destruction of brain areas that are responsible for memory and attention. Also human data of Flöel and colleagues have shown that calorie restriction positively affects age-related declines in cognitive abilities.

Gaining information via basic understanding of the (hormonal) regulatory mechanisms of muscle mass and muscle function is currently essential to improve existing strategies for weight loss as well as to define new therapeutical interventions for sustained body weight reduction. The analysis of these regulatory mechanisms constitutes the focus of this study.

**4	Overview of Trial Design**

**4.1	Specific hypotheses and aims**

The goal is to conduct a prospective randomized intervention study in overweight and obese adults. Specific aims of this proposal are:

Primary aim:

1. Describing the effects of weight loss on muscle mass, strength and metabolism.

Secondary aims:

2. Describing hormonal and metabolic changes at systemic and muscular level that occur during muscle mass reduction during (a) negative energy balance or (b) stabilized body composition after weight loss.
3. Evaluation of long-term effects of weight loss on muscle mass and metabolism in patients regaining weight over time.
4. Evaluation of any metabolic programming of muscle cells ex vivo before and after weight loss.
5. Outlining the changes of the colonic microbiome and virome before and during (a) a period of negative energy balance and (b) a period of stabilized body composition after weight loss and (c) after a period of elongated weight maintenance or after regaining of weight.

**4.2	General study design**

After initial clinical evaluation, at least 80 post-menopausal women will be randomized for an intervention and a control group to enter a 12-weeks controlled randomized life style intervention study according to a randomization list.

Participants of the intervention group will undergo a weight reduction intervention that will be implemented by lifestyle changes by means of recommending increased physical activity and nutrition counseling. In addition, weight loss will be supported by placing study participants on an 800 kcal/d liquid diet during the first 8 weeks of intervention excluding any other food for this time period. This weight loss paradigm will be implemented in analogy to the already approved study „Hormonal regulatory mechanisms in the obese adult after weight loss" (In the context of application for a clinical research group (DFG) KFO 218/0: Hormonal regulation of body weight maintenance). After this initial period, diets will be switched to low calorie regular diets for another 4 weeks to further promote weight loss with healthy foods.

The weight loss period will be followed by 4 weeks of weight maintenance. Here, the diet will be switched to an isocaloric healthy diet that will be supervised by individual nutrition diaries. During this phase, a negative energy balance should be avoided. The increased physical activity will be maintained during this period and weekly nutritional counseling will be continued.

The control group will receive brochures with information about a healthy lifestyle. A further intervention will not take place in this group. Since this study does not target patients with high risk for cardiovascular diseases the study approach seems ethically acceptable.

The main phenotyping of participants in both study groups is scheduled for three study time points: month 0 (M0), month 3 (M3) and month 4 (M4). Additional phenotyping with limited phenotyping parameters is scheduled in the follow-up period after 12 and 24 months. Anticipated weight curves and study phases are depicted in Fig. 1. A study flow diagram is given in Fig. 2.

**Fig. 1	Scheme of study course**

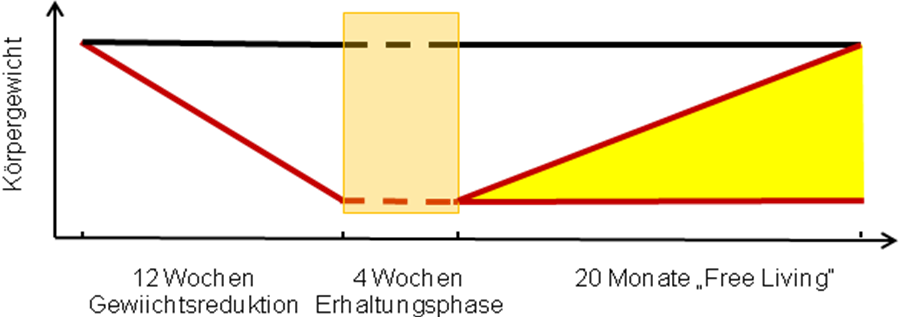

12 weeks weight loss period

4 weeks weight maintenance

20 months „free living“ ad libitum period

Anticipated weight curves

M0

M3

M4

M12

M24

**Time points**

M0 Start of the weight loss period

M3 Start of the weight maintenance period

M4 Start of the free living follow-up period

M12 Follow-up phenotyping after 12 months

M24 Follow-up phenotyping after 24 months

**Fig. 2	Study Flow**

**Screening**

**Informed consent**

**Randomization**

**Phenotyping (M0)**

**Intervention or control group**

**12 weeks weight loss phase (Intervention)**

**12 weeks weight maintenance (Control)**

**Phenotyping (M3)**

**Intervention and control group**

**4 weeks weight maintenance in both groups**

**Phenotyping (M4)**

**Intervention and control group**

**Follow-up: 8 months "free living"**

**Reduced Phenotyping (M12)**

**Follow-up: 12 months "free living"**

**Final reduced Phenotyping (M24)**

**5	Interventions**

**5.1	Weight loss phase**

The protocol of the 12 weeks weight loss program will include caloric restriction, nutritional counseling and motivation for physical exercise. Therefore, weekly meetings will be performed in groups. Caloric restriction will be applied during the weight reduction program in two stages. The first stage will be based on a replacement of all daily meals by a very-low energy diet (Optifast 2^®^, Nestlé HealthCare Nutrition GmbH, Frankfurt am Main, Germany) comprising 800 kcal/d for eight weeks. Each participant will receive 35 portions of formula diet for each week (five per day) within the weekly meetings. All participants will be advised by the nutritionist not to consume any additional foods. After eight weeks of very-low energy diet, the diet will be switched to an energy-reduced healthy diet to facilitate further weight loss. Therefore a balanced mix with the following distribution of macronutrients: carbohydrates 35-45 %, fat 25-35 %, and protein 25-30 % as well as a daily calorie intake of approximately 1500 kcal will be recommended by the nutritionist during this phase. The exact caloric content of this diet will be adapted according to the resting energy expenditure measured during the initial phenotyping (M0) via indirect calorimetry.

This dietary approach will be supported by weekly meetings during the entire weight loss period of 12 weeks. This will also include dietary advices for healthy living and recommendations regarding increased physical activity. During these meetings, nutrition consultants will also perform group workshops with practical cooking exercises. Compliance of the diet will be supported by providing specific recipes, cooking advices, and instructions for behavior modifications. This will focus on only three meals per day, at least 4 hours break between the meals and reduced carbohydrate intake at dinner.

During this phase the control group will be instructed to maintain body weight. Brochures with recommendations on how to live a healthy life will be handed out. Study visits for metabolic phenotyping will take place every 4 weeks. Now further instructions are given.

**5.2	Weight maintenance phase**

After 12 weeks of weight loss in the intervention group and 12 weeks of weight maintenance in the control group both groups will be advised to keep their weight stable for another 4 weeks. Participants of the intervention group will continue with weekly meetings and counseled regarding specific energy needs to maintain body weight after weight loss. The control group will be advised to continue weight stability.

**5.3	Follow-up period**

After completion of the 12 weeks weight loss period and 4 weeks weight maintenance (intervention group) or 16 weeks of weight maintenance (control group), all participants will enter the "free-living" follow-up period. In this period no further dietary and life-style modifying advice will be given to the volunteers. Follow-up study visits will take place at 12 months (M12) and 24 months (M24). During these visits a reduced phenotyping scheme (see below) will take place. The study is completed thereafter.

**6	Definition of primary and secondary endpoints**

**6.1	Primary endpoints**

1. Changes of myocellular insulin sensitivity (hyperinsulinemic clamp) during negative energy balance and during stabilized modification of body composition after weight loss (M3 vs M4).
2. Changes of skeletal muscle mass (air displacement plethysmography) during negative energy balance and during stabilized modification of body composition after weight loss (M3 vs. M4).

**6.2	Secondary endpoints**

3. Effects on energy expenditure; Measurement of energy expenditure (kcal/d), postprandial thermogenesis (%) and respiratory coefficient (M0-M4)
4. Effects on myocellular and adipose tissue metabolism and substrate utilization; Measurement of myocellular and adipose metabolism using microdialysis (glycerol (µmol/l), lactate (mmol/l), pyruvate (µmol/l), glucose (mmol/l)) during oral glucose load (180 minutes)
5. Effects on myocellular and adipose tissue mRNA expression Analysis of myocellular and adipose mRNA expression (RNA sequencing) in counts
6. Weight regain; Analysis of body weight regain (BMI; kg/m2) during follow up
7. Fat mass; Analysis of body fat (kg and %) M0-M24
8. Assessment of the human gut microbiome before, during and after weight loss (negative energy balance) and during stabilized modification of body composition 4 weeks after weight loss via 16S rRNA sequencing and/or shotgun metagenomic pyrosequencing of the gut microbiota for composition and gene abundances
9. Investigating associations between insulin sensitivity and immune cell composition during weight loss and genetic sequencing of the T-cell receptor
10. Comparison of metabolic parameters in different subcutaneous fat depots
11. FFA during negative energy balance and during stabilized modification of body composition after weight loss; Measurement of fatty acids at baseline, during negative energy balance, during stabilized modification of body composition after weight loss and during follow up
12. Metanephrines during negative energy balance and during stabilized modification of body composition after weight loss; Measurement of metanephrines at baseline, during negative energy balance, during stabilized modification of body composition after weight loss and during follow up
13. Leptin during negative energy balance and during stabilized modification of body composition after weight loss; Measurement of leptin at baseline, during negative energy balance, during stabilized modification of body composition after weight loss and during follow up
14. Cortisol during negative energy balance and during stabilized modification of body composition after weight loss; Measurement of cortisol at baseline, during negative energy balance, during stabilized modification of body composition after weight loss and during follow up
15. Follistatin during negative energy balance and during stabilized modification of body composition after weight loss; Measurement of follistatin at baseline, during negative energy balance, during stabilized modification of body composition after weight loss and during follow up
16. Adiponectin during negative energy balance and during stabilized modification of body composition after weight loss; Measurement adiponectin at baseline, during negative energy balance, during stabilized modification of body composition after weight loss and during follow up
17. Natriuretic peptide during negative energy balance and during stabilized modification of body composition after weight loss; Measurement of natriuretic peptide at baseline, during negative energy balance, during stabilized modification of body composition after weight loss and during follow up.
18. IGF-1 during negative energy balance and during stabilized modification of body composition after weight loss; Measurement of IGF-1 at baseline, during negative energy balance, during stabilized modification of body composition after weight loss and during follow up
19. Analysis of the predictive impact of several hormonal and metabolic parameters on body weight regain, course of insulin sensitivity and metabolism; The effect of measured parameters (see other endpoints) on long-term course of BMI, muscle mass, insulin sensitivity and energy expenditure will be analyzed using mathematical models

**7	Study population**

Post-menopausal women will be recruited to participate in this randomized life-style intervention study.

**7.1.	Inclusion criteria**

- BMI > 27 kg/m^2^
- Age > 18 years
- postmenopausal state

**7.2	Exclusion criteria**

- weight loss of more than 5kg in the last 2 months
- unhealthy patients with: severe chronic diseases including cancer within the last 5 years, severe heart disease, severe impairment of hepatic or renal function, severe anaemia or disturbed coagulation
- eating disorders or any other psychiatric condition that would interact with the trial intervention
- malabsorption
- acute or chronic infections
- severe hypertension
- myopathy
- food allergies
- any other uncontrolled endocrine disorder
- changes of smoking habits, diets or medication that strongly affects energy homeostasis within the last 3 months prior to study inclusion

**7.3	Recruitment**

Volunteers will be recruited via leaflets, the outpatient clinic of the Clinic for Endocrinology, Diabetes and Metabolic Diseases and newspaper advertisement.

**7.4	Screening**

Interested candidates will first be screened via phone. The phone interview will be designed to exclude individuals who are clearly ineligible or unlikely to benefit from the study. All potentially eligible individuals will be screened during a clinic visit. Some individuals who visit our outpatient clinic and express interests in participating may be screened on site without a prior phone call. During the screening clinic visit, medical and functional exclusion criteria will be assessed in detail. This includes assessment of medical history and physical examination. Screening will be particularly performed to rule out abnormal thyroid function and hypercortisolism. Therefore TSH levels and 1 mg dexamethasone suppression tests will be performed. Moreover, eGFR, liver enzymes, sodium, potassium, calcium as well as lipid profile (LDL, HDL, total cholesterol and triacylglycerol) will be measured.

**8	Measures and Procedures**

**8.1	Informed Consent**

**Approval from ethical committee for human studies**

The study protocol was approved by the Institutional Review Board of the Charité Medical

School and only subjects, who give written informed consent prior to inclusion in the study,

will be included in the trial.

Approval Number: EA20100415

Board Name: Institutional Review Board of the Charité Medical School

Board Affiliation: Institutional Review Board of the Charité Medical School

Phone: 030/450-517222

Address:

Dr. med. Katja Orzechowski

Charité - Universitätsmedizin Berlin

Ethikkommission

GESCHÄFTSSTELLE

Schumannstr. 20/21

10117 Berlin

**Approval for animal studies**

Does not apply

**8.2	Procedures**

**8.2.1	Phenotyping**

Phenotyping visits will take place at baseline (M0), after 3 months weight loss phase or weight stability (M3) and after 1 month weight stability (M4), as well as after 12 (M12) and 24 (M24) months during the follow-up phase with a reduced set of phenotyping measurements. Participants of the intervention group will additionally visit the study center once per week to attend counseling sessions, undergo basic anthropometrics like body weight measurements and provide stool samples. Additionally, all participants will be asked to answer questionnaires regarding life style, food preferences and intake, mood, physical activity and prior diseases during the main study visits.

Phenotyping at M0, M3 and M4 includes:

- Anthropometry (bioimpedance and body plethysmography measurements to estimate body composition, weight, height [BMI], waist, hip and blood pressure measurements)
- Stool sampling
- 24-h urine sampling (cortisol and metanephrines)
- Oral glucose tolerance test
- 3-day nutrition protocol
- Questionnaires on individual circadian rhythm (Munich Chronotype Questionnaire)
- Indirect calorimetry via a ventilatory hood system and in a metabolic chamber
- Biopsies of adipose and muscle tissue
- Bio-sampling (blood, saliva)
- Euglycemic hyperinsulinemic glucose clamp
- Functional magnetic resonance imaging of the brain in a subgroup of the study population
- Tissue microdialysis (fat and muscle) for measurements of in vivo lipolysis, glycolysis and perfusion during an oral glucose tolerance test in a subgroup of the study population

To integrate this in depth metabolic phenotyping, each phenotyping timepoint will take place during 5 consecutive days.

**8.2.2	Anthropometry**

Height and weight (for BMI); waist, hip, blood pressure and heart rate will be evaluated. Weight will additionally be evaluated in the intervention group during weekly counseling sessions.

**8.2.3	Body composition**

Body composition will be assessed in the fasting state (after 12 h overnight fasting). Subjects are placed in recumbent position for 20 min. Bioelectric impedance measurements using AKERN BIA 101 (SMT medical GmbH & Co. KG, Würzburg, Germany) will take pleace to estimate body composition (fat and fat free mass).

Additionally, whole body plethysmography will be applied using the COSMED BodPod System, Rome, Italy). For this, volunteers will enter a closed chamber in underwear wearing a swim cap. Changes in chamber pressure are then applied to measure whole body volume and estimate body composition.

**8.2.4	Stool sampling**

Stool sample collection tubes and a collection kit as well as thermal packs will be provided. Volunteers will be asked to provide at least two stool samples during the end of a 3-day nutrition diary. Volunteers will be advised to immediately store samples at -20 C in their home freezer in opaque bags and bring samples to the study center during study visits using the thermal packs to guarantee cooling chain. Samples will be immediately transferred to a -80 C study freezer. Sampling will take place in both study groups during the main study visits and during the counseling meetings in the intervention group.

**8.2.5	3-day nutrition diary**

Prior to each study visit volunteers will be asked to fill in a food diary for three days (with at least one weekend day) and document all foods and liquids consumed during the period.

**8.2.6	Social, economic and health related questionnaires**

For descriptive purposes, the following participant characteristics will be collected: age, race, living situation, household composition, marital status, educational level, income, smoking status, alcohol consumed, employment status. Moreover, medical history and family history will be assessed.

The following specific questionnaires will be also used:

International Physical Activity Questionnaire (IPAQ)

SF-36

Eating Behavior Inventory (EBI)

Munich Chronotype Questionnaire

**8.2.7	24-hour-urine sampling**

Volunteers will collect urine samples over 24 h at 3 time points during the study course. Since urine samples need be to mixed with hydrochloric acid in order to stabilize catecholamines (10 ml of 25% hydochloric acid in one 3 l container), there is risk for skin irritation and injury if handled improperly. In this regard, subjects will be comprehensively informed during the informed consent including proper handling of urine sample containers. Additional 24-hour-urine sampling for glucocorticoid assessment will take place on a separate day.

**8.2.8	Oral glucose tolerance test**

An oral glucose tolerance test (oGTT) will be performed at 8 a.m. after 12 h of food restriction by application of 75 g glucose followed by blood sampling over 3 h (glucose and insulin determination at 0‘, 30‘, 60‘, 90‘, 120‘ and 180‘). These values allow the characterization of glucose metabolism as well as the evaluation of the basal insulin sensitivity by application of the HOMA and OGIS models.

**8.2.9	Euglycemic hyperinsulinemic glucose clamp**

First, blood samples for the determination of basal insulin and c-peptide levels will be taken from fasted subjects at -30, -20, -10, 0 min. Blood glucose levels will be measured every 5 minutes in a drop of blood from the earlobe using the glucose oxidase reaction. At t=0 min, subjects will receive an insulin bolus injection followed by an insulin infusion treatment on a constant rate (40 mIU/m^2^/min human insulin, Actrapid®, Novo Nordisk, Bagsvaard, Denmark). Here, blood glucose levels will be adjusted to 4.5 mM by variable glucose infusion (10% glucose solution) and the target blood glucose level will be kept in equilibrium over 3 hours. Insulin concentrations and glucose consumption rates will be measured over time and will be applied for calculating insulin sensitivity.

**8.2.10	Indirect calorimetry**

Following a 10-hour overnight fast, resting energy expenditure will be estimated using indirect calorimetry via a ventilatory hood system (Vmax ENCORE, CareFusion Germany 234 GmbH, Germany) after a 20 minute resting period in recumbent position at 8.00 a.m. Additionally, measurement of total energy consumption will be performed inside a metabolic chamber, which is continuously flowed through by a constant rate of air. The exhaled air will be analysed for the decreasing amounts of oxygen and the total release of carbon dioxide over time. Both values provide a measure for the current metabolic state. Here, where appropriate the examination will be performed under physical exercise (see spiroergometry). After 3 hours fasting in the chamber, volunteers will consume a standardized meal with a caloric content adjusted to each resting metabolic rate and subsequent changes in energy expenditure and respiratory quotient will be monitored for 5 h to measure prandial thermogenesis.

**8.2.11	Fat and muscle biopsies**

Tissue biopsies of abdominal subcutaneous adipose tissue and of the musculus gastrocnemius will be obtained under local anesthesia with a semi-automatic biopsy device (CR Bard GmbH, Karlsruhe, Germany). For this purpose, a local anesthetic with 2 % lidocaine will be applied about 10 cm laterally of the belly button or in the area of the calf followed by a minimal skin incision (3-5 mm) in the anesthetized area with a scalpel. Then a 12G biopsy needle is inserted and by means of high-speed biopsy (very fast automatic pulling back and forth of the biopsy needle) a small sample of muscle or adipose tissue (approx. 20 mg or 200 mg, respectively) is removed and will be immediately freezed into liquid nitrogen. This procedure will be repeated up to 20 times in order to obtain an adequate amount of adipose tissue. The biopsy sampling of muscle tissue will be performed 2 to 3 times. Sampling will be always performed via the already existing skin incision. In some cases, muscle and adipose tissue biopsies will be removed by using a Bergström-Needel. In general, the sampling in the area of the abdominal adipose tissue will be conducted with a Bergström-Needle or a custom made needle as previously published (18) as well according to the procedure described above.

Muscle biopsies will be sampled at the site of the M. quadriceps. Therefore, a local anesthetic with 2 % lidocaine will be applied 15 to 20 cm above the knee joint. Following this, a minimal skin incision (3-5 mm) will be set with a scalpel in the anesthetized area before a needle (4-5 mm outside diameter) will be inserted with a closed biopsy needle. Then, after advancing the cutting cylinder the inserted tissue is will be clipped. Here, the participant might feel a short, dull or stinging pain. After tissue sampling the needle will be removed immediately. The described procedure will only take a few seconds and may be repeated several times depending on the sample mass being recovered. Here, the needle is inserted through the same skin incision, but in a slightly different direction and angle, in order to obtain samples from different sites of the muscle. The further procedure does not differ from the biopsy sampling carried out so far. After completion, which overall will last only a few minutes, pressure will be applied to the wound for approx. 5-10 min to prevent bleeding and hematoma. Thereafter, the skin edges will be closed by Steristrips and will be covered by further bandaging. For the following 12 hours, a slightly compressing bandage with elastic bandages will be applied to counteract bruising. Subjects will be advised to abstain from sports for 24 hours, although general physical activity is unproblematic.

In addition to the biopsies of adipose tissue from the abdominal subcutaneous fat, biopsies from subcutaneous fat of the lateral thigh / gluteal region will be obtained as well in a subgroup. The procedure will be analogous to the biopsy procedure of the adipose tissue from the abdominal subcutaneous fat.

Biopsies will be used for mRNA analysis, measurements with the Seahorse XF-Analyzer (ECRC-MDC) as well as for detection of metabolites (Max Planck-Institute of Molecular Plant Physiology).

**8.2.12	Blood sampling and salivary cortisol**

Blood will be screened for any severe diseases and general metabolic parameters (HbA1c, hormone marker, cytokines) by a clinical standard laboratory panel. In addition, an aliquot will be applied for a functional analysis of monocytic cells on mRNA level (Max-Delbrueck-Zentrum für Molekulare Medizin, Berlin-Buch). No genetic analysis will be applied. The circadian cortisol levels will be determined by measuring the cortisol concentrations in saliva samples at different times of day. Here, patients will repeatedly collect saliva samples in special salivary tubes (salivettes from Sarstedt) within 24 hours. The test is a commonly used, fast and hygienic procedure for the analysis of cortisol in saliva.

**8.2.13	Functional magnetic resonance imaging**

In a subgroup of the study population, a fMRI examination of the head will enable the determination of gray matter function in the context of central nervous information processing. In doing so, the principle of “neurovascular coupling" is exploited: activation of an area of the brain goes along with an increased perfusion and thus an increase in the proportion of oxygenated hemoglobin. Differences in signal measurements of oxygenated versus deoxygenated hemoglobin allows the indirect analysis of focal neuronal activation states under standardized stimulation conditions and further allows conclusions about the extent and intensity of central nervous activation states of the gray matter. The paradigm of cue-reactivity constitutes a standard paradigm adopted from addiction research to evoke activity of the fronto-striatal compounds. This paradigm used to investigate cue reactivity has been tested in a preliminary study. Here, 30 images (foods / dishes) out of three categories (high calorie, low calorie and food related) are presented to our volunteers as well as a block of 10 neutral images (e.g., flowers, field). The different blocks are presented several times in alternating order. Subjects will be asked to attentively view the images, further reactions or actions are not necessary. The total duration for this experiment is about 30 minutes. The paradigm of reward delay is a modified approach. In the present study, subjects are repeatedly given the choice of a direct (on the day of the experiment) but low reward or of a delayed (one month after the experiment), but more extensive reward in the form of their favorite chocolate. In this case, the decision to apply idiosyncratic amplifiers was made against the background of psychotherapeutic studies. Thus, it has been shown that appropriate interventions which are based on symptom provocations are all the more successful when based on stimuli from the daily lives of patients. In addition, it is assumed that such stimulus types might generate a maximal decision conflict and thereby evoke corresponding activation effects. The precise extent of the immediate and delayed reward and thus their relation to each other will be determined in advance by means of a preliminary experiment in which in a small subcohort of the subjects on the basis of maximum reaction time differences.

The design of the experimental trial is as follows: both choices will be displayed simultaneously on a computer screen. Below each choice a green triangle will be presented. Subsequently, subjects will have unlimited time to decide on one of the two options. As soon as subjects have made a choice by pushing a keyboard button, the triangle under the unselected option will disappear and the one under the selected option will be displayed in red to indicate selection. This display will persist for two seconds. Subsequently, a black screen will be displayed for 12 seconds and the procedure will be repeated for several cycles. The overall duration of the experiment will last 30 minutes, thus the number of passed cycles will result from the individual speed of the subjects. Furthermore, the German version of the “Three-factor Eating Questionnaire” will be collected in order to investigate behavioral aspects such as cognitive restraint, uncontrolled eating and eating for emotional reasons as well as their relation to subcortical and cortical activations and deactivations, which might facilitate the usage of these behavioral patterns for the prediction of therapeutic success by the pattern recognition method. The German version of the “Perceived Stress Questionnaire” will be applied to investigate the extent to which the currently perceived stress affects the neuronal activation. Moreover, the Self Assessment Manikin (SAM) will be applied to determine valence, arousal and dominance of the images that were used in the cue reactivity paradigm. All questionnaires will be collected at all three timepoints.

In addition, besides a T1-weighted 3D sequence with an isotropic resolution of 1 x 1 x 1 mm (or 0.5 x 0.5 x 0.5 mm), an axial FLAIR and a DTI sequence (so-called diffusion tensor imaging) will be measured. Moreover, a functional sequence for measuring the BOLD signal will be performed at rest (resting-state connectivity). The MR images will be evaluated by various methods, i.a. "Voxel-Based-Morphometry" and "Tract-Based-Spatial Statistics”, using several software packages (SPM and Matlab, FSL and AFNI).

These studies constitute surrogate markers for structural / functional neuronal plasticity. The additional examinations might cause a prolonged measuring time in the MRI (3 Tesla MRI scanners (Siemens)) of approx. 20 minutes, the whole examination time will take about 50 minutes.

**8.2.14	Subcutaneus microdialysis**

Microdialysis of adipose tissue will be performed at timepoint M0, M3 and M4. Microdialysis allows metabolic and hemodynamic assessment, which can be carried out in different tissues simultaneously. Therefore, a microdialysis catheter (CMA 60, Sweden) will be inserted into the subcutanous abdominal adipose tissue and the thigh muscle. Due to the concentration gradient between interstitial fluid and perfusate, either substrates / effectors or metabolites diffuse into the tissue or the perfusate, respectively. Effects on hemodynamics (blood circulation) in the tissue can be detected by means of the so-called ethanol dilution technique. To this end, a certain concentration of ethanol is added to the perfusate (50 mM) and the local circulation can be quantified by the ratio of ethanol concentration in the dialysate and ethanol concentration in the perfusate. The protocol takes about 4 hours, plus about 60 min. of pre- and post-processing. Finally, the *in vivo* -activity of the 11β-hydroxysteroid dehydrogenase type 1 will be evaluated by measuring the free cortisol by RIA in the microdialysate.

**8.3	Assessment Schedule**

On overview of the main clinic visits, procedures and assessments is given in Table 1. Volunteers in the intervention group additionally attend weekly counseling sessions.

Table 1: Assessments schedule

| Study visits | M0 | | M3 | | M4 | | M12 | M24 |
| --- | --- | --- | --- | --- | --- | --- | --- | --- |
| Anthropometry | X | | X | | X | | X | X |
| Body Composition | X | | X | | X | | X | X |
| Stool Sampling | X | | X | | X | | X | X |
| 3-Day Nutrition Diary | X | | X | | X | | X | X |
| Questionnaires | X | | X | | X | | X | X |
| 24 h Urine Sampling | X | | X | | X | |  |  |
| Oral Glucose Tolerance Test | X | | X | | X | | X | X |
| Euglycemic Hyperinsulinemic Clamp | X | | X | | X | |  |  |
| Indirect Calorimetry | X | | X | | X | |  |  |
| Fat and Muscle Biopsies | X | | X | | X | |  |  |
| Blood sampling and salivary cortisol | X | | X | | X | | X | X |
| Functional magnetic resonance imaging | X | | X | | X | |  |  |
| Subcutaneus microdialysis | X | X | | X | |  | |  |

**8.4	Randomization**

A stratified randomization of the participants will be performed using a stratified randomization list. Stratification will consider BMI at baseline (3 BMI strata).

**8.5	Participant Safety**

Risks associated with a weight reduction will be assessed. Therefore, fat metabolism (i.e. increase of triglycerides), renal function and electrolytes will be analyzed at baseline, after 4 and 8 weeks during the initial phase of the very low calorie diets at frequent intervals. Body weight will be measured weekly during the 12-week weight loss period. In the first two months weight loss and health status will be monitored weekly, which included a patient interview and blood pressure measurement. Body weight will be measured at least once per week. As a precaution, subjects will be regularly instructed to ensure sufficient fluid intake. The formula diet phase will be conducted with a commercially available liquid diet (commercial formula diet) that contains all the essential micronutrients in daily required dosing. The low caloric intake may initially cause mild discomforts such as fatigue, dizziness, nausea, reduction in stool volume, constipation or stomach problems.

Summary of risks associated with the study: in the context of blood sampling, malpuncture of the vein may occur, and a so-called "blue mark" (bruise) might arise. Very rarely it can also lead to a nerve injury. Furthermore, an infection may develop. In the context of the oGTT no serious side effects are known. Rarely, reactive hypoglycemia may occur. However, this would be noticed during the investigation due to the relatively long observation period. In this case, the participant will be provided with complex carbohydrates (e. g. whole wheat bread). For clamp analysis, blood sampling and measurements may be implemented by 2 indwellling intravenous catheters. There is a low risk of a thrombophlebitis and a low risk of malpuncture. A potential inflammation of the venes will be treated by anti-inflammatories. For the case of malpunction, the affected vessel will be manually compressed and a follow-up supervision will take place. The volume of blood sampled is usually unproblematic in healthy volunteers. The most common side effect is the drop in blood glucose by the prolonged action of insulin. For this reason, the test subject will be further monitored for about 1 hour and the glucose infusion rate will be gradually phased out. For urine sampling there is a risk for skin irritation and injury if samples will be handled improperly. In this regard, the subjects will be comprehensively informed within the declaration of consent including the proper handling of urine sample containers. Risks of biopsy sampling comprise potential infection and hemorrhages. If there is no complication, a small pigment shift might occur. The risk of infection will be minimized by aseptic work, bleeding will be treated by compression. The occurrence of reversible, uncomplicated hematomas can not be generally excluded and will be treated conservatively with cooling devices and, if necessary, with ointment bandaging. Heavier hemorrhages / complications can not be entirely excluded. If necessary, they will be treated according to the medical requirements. In order to detect and treat possible complications (e. g. infection, bleeding, puncture of the abdominal cavity) in a good manner, patients will be invited to present their complaints at any time in our department. An existing hypersensitivity to anesthetics will be excluded by medical history as far as possible. The risks of microdialysis are low. Very rarely, infections and bleeding may occur. Heavier bleeding is extremely unlikely. However, no complications have occurred within the scope of previous microdialysis. There will be no radiation exposure during magnetic resonance imaging, and so far no other detrimental side effects or long-term side effects of magnetic resonance imaging have become known. Thus, magnetic resonance imaging is harmless for patients. magnetic resonance imaging cannot be applied to subjects with pacemakers due to magnetic field interference. The presence of electrically, magnetically or mechanically activated metallic implants is generally an exclusion criterion. These include piercings or tattoos with metallic colors. Whether patients with metal implants (e.g. a ferromagnetic aneurysm clip) or with movable metal prostheses can be examined requires prior clarification in individual cases. Participants who suffer from panic and anxiety in confined spaces (claustrophobia) will be excluded from this investigation as well. Imaging by MRI will be performed in accordance with the Medical Device Act and Medical Device Supply. For this purpose, patients will be informed in detail and the presence of possible metallic implants will be queried beforehand. Considering the contraindications, magnetic resonance imaging scans might rarely lead to mild side effects (headache or exposure to loud noise from the scanner). Basal metabolic rate will be determined by measuring the respiratory coefficient. This is a routine clinical examination, however resting under the ventilatory hood and in the closed metabolic chamber may cause initial discomfort.

A Data Safety Monitoring Board (DSMB) will be established, with responsibility to monitor all safety aspects of the study. The Medical Safety Committee reports to the DSMB for issues related to participants safety. The monitoring will be provided by the KKS of the Charité.

A volunteer will be excluded from the trial at the following conditions:

 the user's own request (also without a reason)

 if considered necessary by the investigator or head of the study from a medical point of view (e.g., due to abnormal laboratory values)

 the subject does not follow the study protocol (e.g. substantial deviation from the recommended diet)

 severe disease

 new scientific findings during the study's lifetime, which are against the continuation

of the clinical trial

**9	Data management**

Data collection will be performed using CRF. All data will be transferred by independent staff into an electronic data base.

**10	Statistical Considerations**

**10.1	Power analysis and sample size calculation**

Previous studies in adults report that a weight reduction by a change in lifestyle over 3 months is accompanied by a reduction in muscle mass from 3.2 to 6.9 %. Assuming an effect of at least 3.2% and a variance of 3.4%, the power calculation yields a case number of 19 subjects per treatment arm (significance level 0.05 two-sided, power 80%, nquery 7.0). This effect difference refers to the end of the study intervention (maintenance phase 4 weeks after the end of the weight reduction program, T4). With an expected drop out number of 25 % during the weight loss phase and 15 % in the 4-week maintenance period, respectively, a total of at least 30 adults per treatment group should be included in the trial at the time of randomization.

**10.2	Data analysis plan**

Study outcomes will be analyzed by an intention to treat (ITT) analysis as well as a per-protocol analysis. The intention-to-treat (ITT) population is defined by all randomized patients who have participated in at least one study visit.

Mixed-model, repeated-measures analysis of variance will be used to analyze the primary outcomes. This will consider the correlation between repeated observations and will use all available subsequent observations for all participants with values at randomization, regardless of further assessment completion. Means/Medians will be modeled as a function of group assignment and study visit (M0, M3, M4). Additional adjustment for age and BMI before weight loss will be included in this model if necessary. Secondary endpoints will mostly be analysed in a PP approach to make use of all data available. This will include mixed model, repeated-measures analysis of variance to compare hormonal levels of intervention and control group within the trial. Adjustment for potential confounders like age and BMI will be included in these models. Prediction analyses will be performed using linear or logistic regression models.

Multidimensional data deriving e.g. from gene sequencing will be analysed by methods mentioned above and additionally handled using machine learning methods.

**11	Appendix**
